# Mapping brain structural differences and neuroreceptor correlates in Parkinson’s disease visual hallucinations: a mega-analysis

**DOI:** 10.1101/2021.02.17.21251558

**Authors:** Miriam Vignando, Dominic ffytche, Simon Lewis, Phil Hyu Lee, Seok Jong Chung, Rimona S. Weil, Michele T. Hu, Clare E. Mackay, Ludovica Griffanti, Delphine Pins, Kathy Dujardin, Renaud Jardri, John-Paul Taylor, Michael Firbank, Grainne McAlonan, Henry Ka-Fung Mak, Shu Leong Ho, Mitul A Mehta

## Abstract

*Parkinson’s psychosis* (PDP) describes a spectrum of symptoms that may arise in Parkinson’s disease (PD) including visual hallucinations (VH). Imaging studies investigating the neural correlates of PDP have been inconsistent in their findings, due to differences in study design and limitations of scale. Here we use empirical Bayes harmonisation to pool together structural imaging data from multiple research groups into a large-scale mega-analysis, allowing us to apply new methodological approaches to identify cortical regions and networks involved in VH and their relation to receptor binding. Differences of cortical thickness and surface area show a wider cortical involvement underlying VH than previously recognised, including primary visual cortex and its surrounds, and the hippocampus, independent of its role in cognitive decline. Structural covariance analyses point to a strong involvement of the attentional control networks in PD-VH, while associations with receptor density maps suggest neurotransmitter loss may drive the cortical changes.

## 1. Introduction

Parkinson’s disease (PD) is a neurodegenerative disorder primarily characterised by motor symptoms, mainly related to the loss of neurons in the substantia nigra projecting to the basal ganglia (Dickson et al., 2009). Patients with PD commonly experience a variety of non-motor symptoms, including psychiatric ones (Schapira et al., 2017). Among these, visual hallucinations (VH) and related visual phenomena form a spectrum of symptoms referred to as *Parkinson’s psychosis* (Ravina et al., 2007) (PDP). There is a continuum of experiences typically characterising PDP with patients initially experiencing minor hallucinations (perception of presence or passage) and illusions that progress to formed hallucinations (initially with insight preserved), then hallucinations in other modalities and delusions (ffytche et al. 2017). Such symptoms may affect up to 70% of PD patients in more advanced stages of the illness (Levin et al., 2016) in the context of dopamine therapy but do not show a clear relationship between medication introduction or dose suggesting they are not simply medication side-effects (ffytche et al. 2017). VH predict a range of poor outcomes including more rapid cognitive decline and development of dementia (Aarsland et al., 2003; Anang et al., 2014; Uc et al., 2009) and increased likelihood of a move from independent living to a care home (Goetz and Stebbins, 1993; Aarsland et al., 2000). It is difficult to determine how VH might be related to these poor outcomes without a clear understanding of the brain systems involved in VH (ffytche et al., 2017).

Imaging studies of VH in PD to date have been based on relatively small samples and have used differing designs that variously control for the degree of cognitive decline, stage of PD and dopamine medication. This makes it difficult to disentangle brain changes related specifically to VH mechanisms as distinct from those related to cognitive decline, PD stage or medication effects. As a result, a heterogeneous array of structural differences has been reported. Depending on whether or not cognition is controlled for, some studies have found volume reductions in specific regions that have not been replicated in other studies including: hippocampus (Ibarrexete-Bilbao et al., 2008), cerebellum (Ibarrexete-Bilbao et al., 2008; Watanabe et al., 2013), lateral, superior and medial frontal cortex (Sanchez-Castaneda et al., 2010; Gama et al., 2014; Watanabe et al., 2013) thalamus (Shin et al., 2012) and different subregions of visual association cortex, broadly defined to include the lateral occipital cortex, ventral occipito-temporal cortex (ventral stream) and visual parietal lobe (dorsal stream) (Goldman et al., 2014; Ramirez-Ruiz et al., 2007; Watanabe et al., 2013).

A meta-analysis (Weil et al., 2019) utilising the previously reported regional differences demonstrated very little consistency across studies. It suggested this may be due to heterogeneity in structural brain correlates of VH, varying sensitivity to detect differences in multiple small studies, or the involvement at different locations of a unifying brain network whose dysfunction results in VH (Weil et al. 2019). While meta-analytical techniques can be useful to collate findings from different studies and help understand the consistency of brain regions involved, there are limitations in their ability to include variables such as cognition, medication dose, PD stage and duration as covariates, given that these are usually incorporated into the analyses at the study level and each study contributes a different set of regions to the meta-analysis. In contrast, mega-analyses bring together subject-level data across sites in one analysis, which presents a number of advantages. These include methodological rigour, with shared quality control and pre-processing pipelines, including software version control and the ability to include unpublished data or published data that was not used in the primary analysis (e.g. structural data collected for functional imaging studies). The same experimental design model and covariates can be applied uniformly across the data set helping address design variations in previous studies. Another advantage of the increased sample size is the additional power to explore morphometric features such as cortical thickness and cortical surface area along with undertaking complex analyses, such as structural covariance. Cortical thickness and surface area are considered as orthogonal components, which are genetically unrelated (Panizzon et al. 2009) and can be considered separate morphometric components in ageing and disease (Dickerson et al. 2009; Storsve et al. 2014). The main correlate of cortical volume is cortical surface area, but volume loss is best captured by cortical thickness (Im et al. 2008; Storsve et al., 2014). Separate measurement and analysis of these two components thus offer a better understanding of the underlying cortical changes associated with VH in PD than volume measures alone. Finally, mega-analyses create a valuable resource that can evolve and be made available to the wider neuroimaging community, especially important in PDP given that such patients are difficult to recruit and scan.

Several neurotransmitter systems have been associated with VH in PD. Initially, VHs were proposed to be a side effect of *dopaminergic* medication (Goodwin, 1971), but later evidence has led to a revision of this view. Current consensus is that dopaminergic medication interacts with disease-related susceptibility factors in PD to cause VH, rather than as a simple side effect (Ravina et al., 2007). Cholinergic pathways have also been implicated in VH (e.g. Janzen et al., 2012; Collerton et al., 2005), with neurodegeneration in brainstem and forebrain cholinergic nuclei (Janzen et al., 2012) and electrophysiological measures of cholinergic function reduced in patients with VH (Manganelli et al. 2009). Recently, a role for serotonergic dysfunction in VH has been suggested (Ballanger et al., 2010), linked to alterations of 5-HT_2A_ receptor density (Yasue et al., 2016; Huot et al, 2010) (for a review Powell et al., 2020).

In summary, our mega-analysis of PD with VH compared to PD without VH enables analyses that are not available to smaller scale studies to help explore the mechanisms of VH. Specifically, we are able to determine the regional cortical thickness and surface area changes associated with VH and relate these morphometric features to measures of symptom severity in a subgroup where finer-grain clinical detail is available. We perform a principal component analysis to identify smaller-scale morphometric differences within a high dimensional set of regions. In addition, we perform an exploratory structural network analysis to highlight associations between regions and clusters of connections linked to VH. Structural covariance allows us to assay covariation of differences in grey matter morphology between different brain structures, providing information on which regions similarly change in thickness or surface area. In order to understand the neurochemical associations of these changes, we also test the hypothesis that structural differences are related to the spatial variation in subtypes of receptors for which high resolution PET atlases are available (dopamine and serotonin).

## 2. Results

### 2.1 Patient characteristics

The final dataset consisted of 493 participants (193 F), of which 135 were PD-VH. Each individual study had matched their participants for age, gender, disease onset, MMSE, UPDRS-III and levodopa equivalent daily dose (LED), except for a study where MMSE score was lower in PD-VH, a study with UPDRS-III scores higher in PD-VH, and two studies where gender was not matched (**Table 1**). We also included the unpublished data in separate ANOVAs to check that groups were matched (**Table 1**), in meta-analyses (**S2**) and in an ANOVA including the whole mega-analysis sample. While the ANOVAs and the meta-analysis demonstrated we have good matching on the criteria, the mega-analysis ANOVA shows that there is a difference of 2.19 years in age [*F* (1,491) = 6.56, *p* = .01] (PD-VH = 67.85, SD = 7.74; 62 F, and PD-noVH = 65.66, SD = 8.71; 131 F) and there is a greater proportion of females in the PD-VH group (**χ**^2^ = 3.585, *p* = .06). Morphometrics were harmonised (**S1**) and we did not find significant differences in total intracranial volume (TIV) [*F*(1,493) = .043, *p* = .84] and total brain volume [*F*(1,493) = 2.488, *p* = .115], but in total gray matter volume [*F*(1,493) = 5.41, *p* = .02] (see **S2**). For a smaller subsample of patients (N=146) we had additional information and a subsample analysis was carried out (see **2.3**).

**Table 1.**
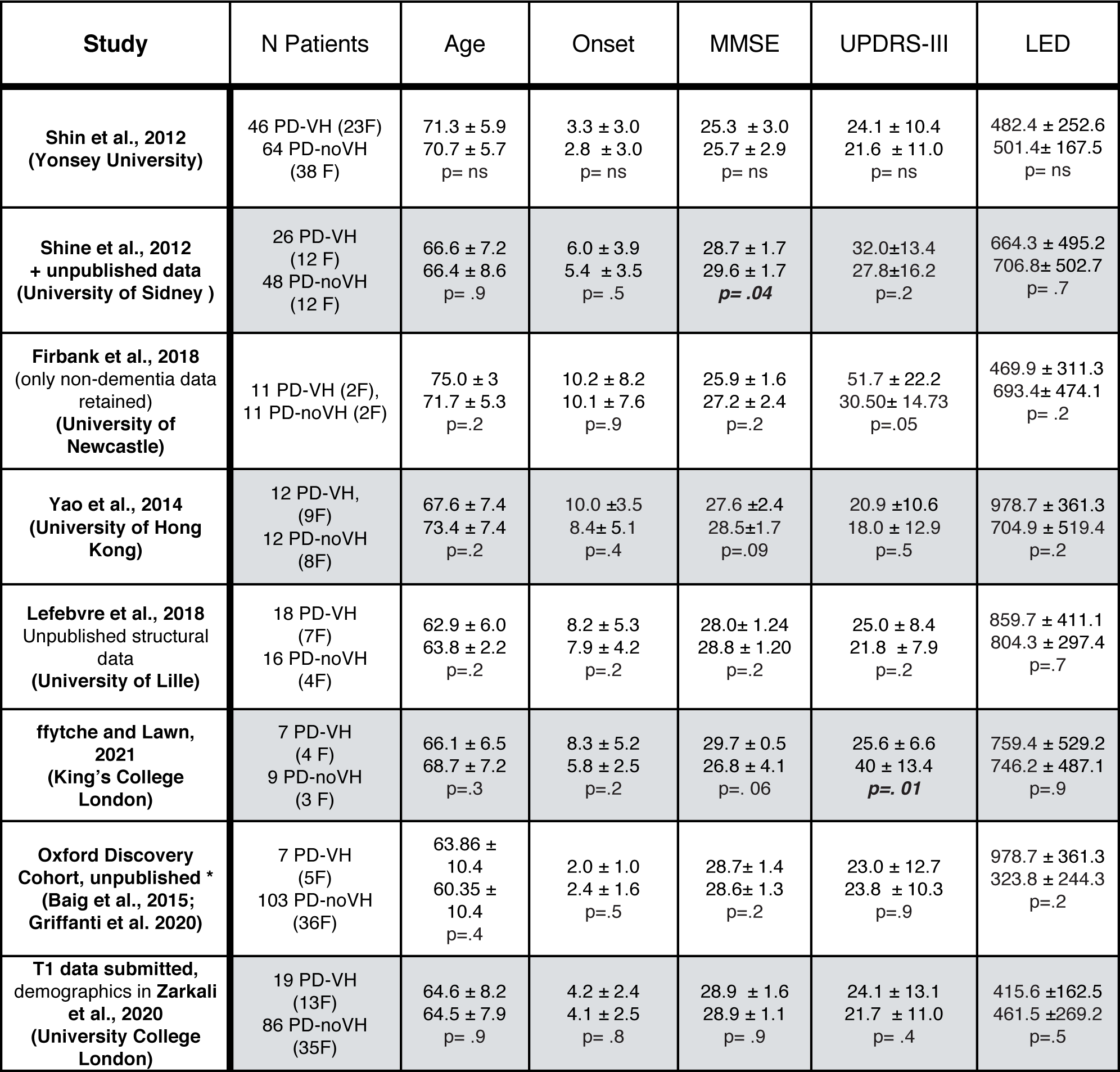
Demographics and clinical information by group. Each row represents the data present in the study for each group. Not all groups could share raw clinical data. In those cases, we reported the information of the original publication to show that there was no difference within groups in terms of PD and medication. Gender was not matched for the UCL and Sidney samples. *\* non-motor symptoms (Baig et al., 2015), T1 data (Griffanti et al., 2020) separately published, but not in a publication studying them together.*

| Study | N Patients | Age | Onset | MMSE | UPDRS-III | LED |
| --- | --- | --- | --- | --- | --- | --- |
| **Shin et al., 2012**<br>(Yonsey University) | 46 PD-VH (23F)<br>64 PD-noVH (38 F) | 71.3 ± 5.9<br>70.7 ± 5.7<br>p= ns | 3.3 ± 3.0<br>2.8 ± 3.0<br>p= ns | 25.3 ± 3.0<br>25.7 ± 2.9<br>p= ns | 24.1 ± 10.4<br>21.6 ± 11.0<br>p= ns | 482.4 ± 252.6<br>501.4 ± 167.5<br>p= ns |
| **Shine et al., 2012**<br>+ unpublished data<br>(University of Sidney ) | 26 PD-VH (12 F)<br>48 PD-noVH (12 F) | 66.6 ± 7.2<br>66.4 ± 8.6<br>p= .9 | 6.0 ± 3.9<br>5.4 ± 3.5<br>p= .5 | 28.7 ± 1.7<br>29.6 ± 1.7<br>**p= .04** | 32.0 ± 13.4<br>27.8 ± 16.2<br>p=.2 | 664.3 ± 495.2<br>706.8 ± 502.7<br>p= .7 |
| **Firbank et al., 2018**<br>(only non-dementia data retained)<br>(University of Newcastle) | 11 PD-VH (2F),<br>11 PD-noVH (2F) | 75.0 ± 3<br>71.7 ± 5.3<br>p=.2 | 10.2 ± 8.2<br>10.1 ± 7.6<br>p=.9 | 25.9 ± 1.6<br>27.2 ± 2.4<br>p=.2 | 51.7 ± 22.2<br>30.50 ± 14.73<br>p=.05 | 469.9 ± 311.3<br>693.4 ± 474.1<br>p= .2 |
| **Yao et al., 2014**<br>(University of Hong Kong) | 12 PD-VH, (9F)<br>12 PD-noVH (8F) | 67.6 ± 7.4<br>73.4 ± 7.4<br>p=.2 | 10.0 ± 3.5<br>8.4 ± 5.1<br>p=.4 | 27.6 ± 2.4<br>28.5 ± 1.7<br>p=.09 | 20.9 ± 10.6<br>18.0 ± 12.9<br>p=.5 | 978.7 ± 361.3<br>704.9 ± 519.4<br>p=.2 |
| **Lefebvre et al., 2018**<br>Unpublished structural data<br>(University of Lille) | 18 PD-VH (7F)<br>16 PD-noVH (4F) | 62.9 ± 6.0<br>63.8 ± 2.2<br>p=.2 | 8.2 ± 5.3<br>7.9 ± 4.2<br>p=.2 | 28.0 ± 1.24<br>28.8 ± 1.20<br>p=.2 | 25.0 ± 8.4<br>21.8 ± 7.9<br>p=.2 | 859.7 ± 411.1<br>804.3 ± 297.4<br>p=.7 |
| **ffytche and Lawn, 2021**<br>(King's College London) | 7 PD-VH (4 F)<br>9 PD-noVH (3 F) | 66.1 ± 6.5<br>68.7 ± 7.2<br>p=.3 | 8.3 ± 5.2<br>5.8 ± 2.5<br>p=.2 | 29.7 ± 0.5<br>26.8 ± 4.1<br>p=. 06 | 25.6 ± 6.6<br>40 ± 13.4<br>**p= .01** | 759.4 ± 529.2<br>746.2 ± 487.1<br>p=.9 |
| **Oxford Discovery Cohort, unpublished \***<br>(Baig et al., 2015; Griffanti et al. 2020) | 7 PD-VH (5F)<br>103 PD-noVH (36F) | 63.86 ± 10.4<br>60.35 ± 10.4<br>p=.4 | 2.0 ± 1.0<br>2.4 ± 1.6<br>p=.5 | 28.7 ± 1.4<br>28.6 ± 1.3<br>p=.2 | 23.0 ± 12.7<br>23.8 ± 10.3<br>p=.9 | 978.7 ± 361.3<br>323.8 ± 244.3<br>p=.2 |
| **T1 data submitted, demographics in Zarkali et al., 2020**<br>(University College London) | 19 PD-VH (13F)<br>86 PD-noVH (35F) | 64.6 ± 8.2<br>64.5 ± 7.9<br>p= .9 | 4.2 ± 2.4<br>4.1 ± 2.5<br>p= .8 | 28.9 ± 1.6<br>28.9 ± 1.1<br>p= .9 | 24.1 ± 13.1<br>21.7 ± 11.0<br>p= .4 | 415.6 ± 162.5<br>461.5 ± 269.2<br>p=.5 |

### 2.2 Hallucinators (PD-VH) vs. non-hallucinators (PD-noVH) multivariate analysis of variance

#### Cortical Thickness

Lower thickness in PD-VH was present in a widespread set of regions (see **Figure 1** and **S3**). No regions showed greater cortical thickness in PD-VH. A main effect of *age* [*F*(1,492)=3.38, *η^2^* = .60, *p* < .001], *gender* [*F*(1,492)=1.51, *η^2^* = .40, *p* <.001] and TIV [*F*(1,492)=2.38, *η^2^* =.51, *p* <.001] was observed.

**Figure 1.**
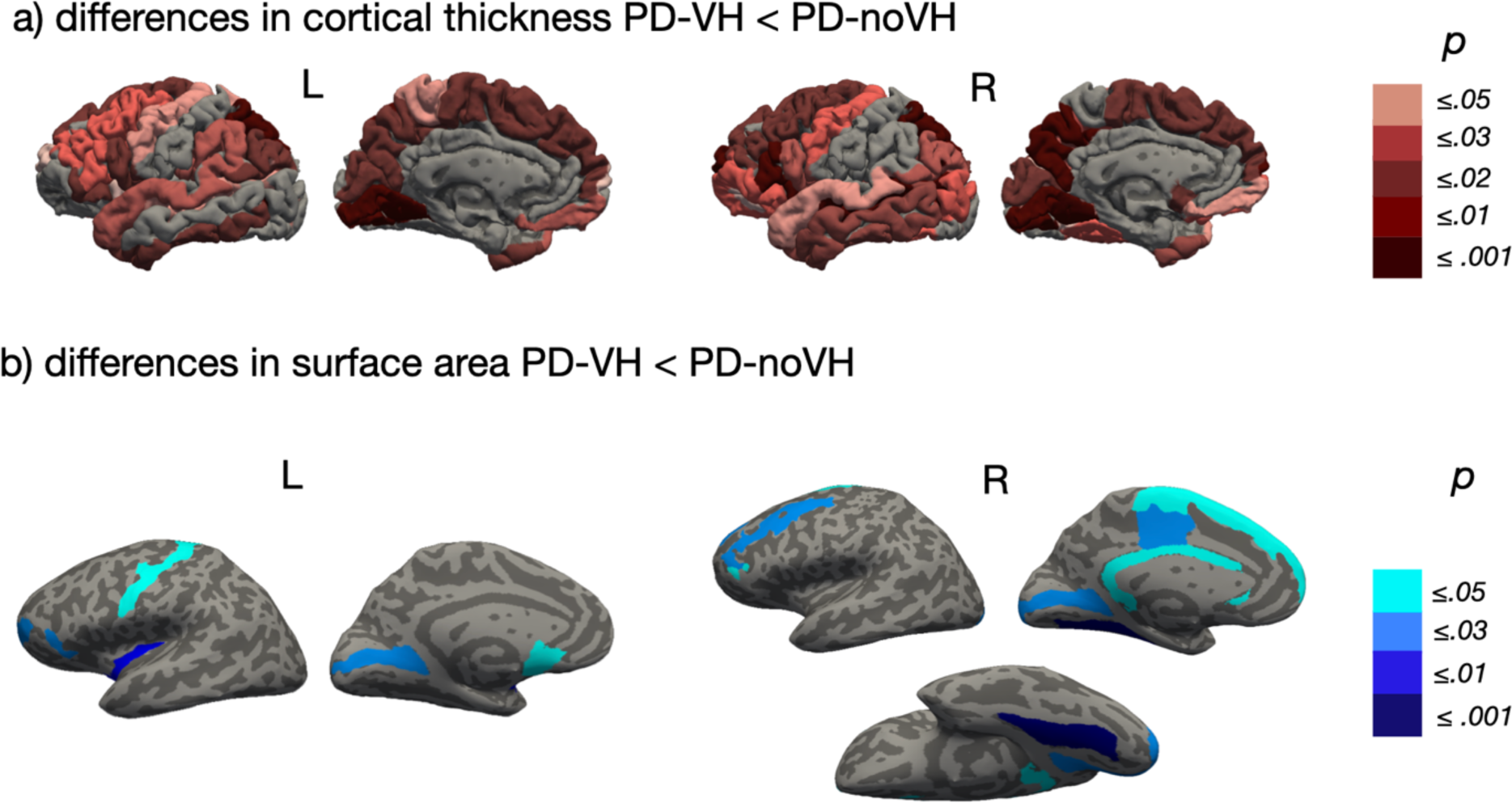
Group differences for PD-VH vs. PD-noVH. Shown are regions whereby PD-noVH had decreased (a) cortical thickness and (b) surface area (SA). Regions are colour coded by p value (**S3**). (a) Widespread decreased thickness was found in PD-VH; the regions with the greatest effect size were medial occipital parietal and frontal regions. (b) SA was reduced in PD-VH in the left and right medial occipital and in the left insular gyrus, and in the medial central and superior frontal regions. Results are corrected for multiple comparisons.

#### Surface area

We found reduced area in PD-VH mainly in frontal and occipital regions (see **Figure 1**) (for all tables and details see **S3**). A significant main effect of *age* [*F*(1,492)=2.08, η^2^ =.47, p <.001], *gender* [*F*(1,492) = 1.50, *η^2^* =.39, *p* <.001] and *TIV* [*F*(1,492) = 6.32, *η*^2^ =.73, *p* <.001] was observed.

#### Subcortical volumes

We found a lower volume for PD-VH in the bilateral amygdala (see **S3**). A significant main effect of *age* [*F*(1,492) = 11.87, *η^2^* = .62, *p* <.001], *gender* [*F*(1,492) = 2.64, *η^2^* = .26, *p* <.001] and TIV [*F*(1,492) = 255.89, *η^2^* = .97, *p* <.001] was observed.

### 2.3 Subgroup analysis

For the subsample for which we have Neuropsychiatric Inventory (NPI) hallucinations subscale scores (*frequency * severity*), focussing on VH, we ran additional correlational analyses. The NPI sample consists of 146 subjects (67 PD-VH, 79 PD-noVH), matched for age, gender, TIV, medication, cognition, onset and PD severity (UPDRS-III) (but see **S4** for detailed comparisons). Results from the custom multivariate ANCOVAs were overall consistent with those found for the main sample (see **S4**).

When correlating the NPI score with morphometrics, inverse correlations were significant for right hemisphere cortical thickness in the intraparietal sulcus (*r* = -.24, *p* = .05), the superior temporal sulcus (*r* = -.26, *p* = .03), the Jensen sulcus (between the anterior and posterior rami of the IPS) (*r* = -.27, *p* = .03) and the cingulum (marginalis) (*r* = -.25, *p* = .05), and a positive correlation was found with the right frontomarginal gyrus (*r* = .26, *p* = .04). Results did not change when carrying out partial correlations between NPI score and morphometrics, and with levodopa equivalent dose (LED) as covariate (see **S4**).

### 2.4 Receptors density maps regression models

After parcellating the receptor densities maps of D2/D3, 5-HT_2A_ and 5-HT_1A_ receptors using the Destrieux atlas to ensure that density and morphometric data were aligned, we explored the relationship between the differences in cortical thickness and surface area between PD-VH and PD-noVH (see **Figure 3**). Separate linear models were carried out for each receptor density map. These correlations were assessed both with a model including the morphometric difference values only of regions where we found a significant difference, and with a model including the morphometric difference values in all regions. The maps used were independent atlases built on healthy subjects’ PET data (see Methods).

#### Thickness

The model with 5-HT_2A_ binding potential as predictor and the mean difference as dependent variable was significant for the subset of regions where the groups differed (*β* =-.252, *t* = -2.2, *p* =.03), whereas no relationship was observed when considering all the atlas regions (*β* =-.02, *t* = -.31, *p* = .75). A similar result was observed for 5-HT_1A_ (significant regions: *β* = -.26, *t* =-2.25, *p* = 0.03; all regions: *β* =.008, *t* =.092, *p* = .504) and for D2/D3 receptors (significant regions: *β* =-.35, *t* =-3.14, *p* = 0.002; all regions: *β* =.09, *t* =.95, *p* =.34) (see **Figure 2** for methods and results). In addition, we compared the slopes of the models, finding no difference between 5-HT_2A_, 5-HT_1A_ and D2/D3 for significant regions) (**S6**).

**Figure 2.**
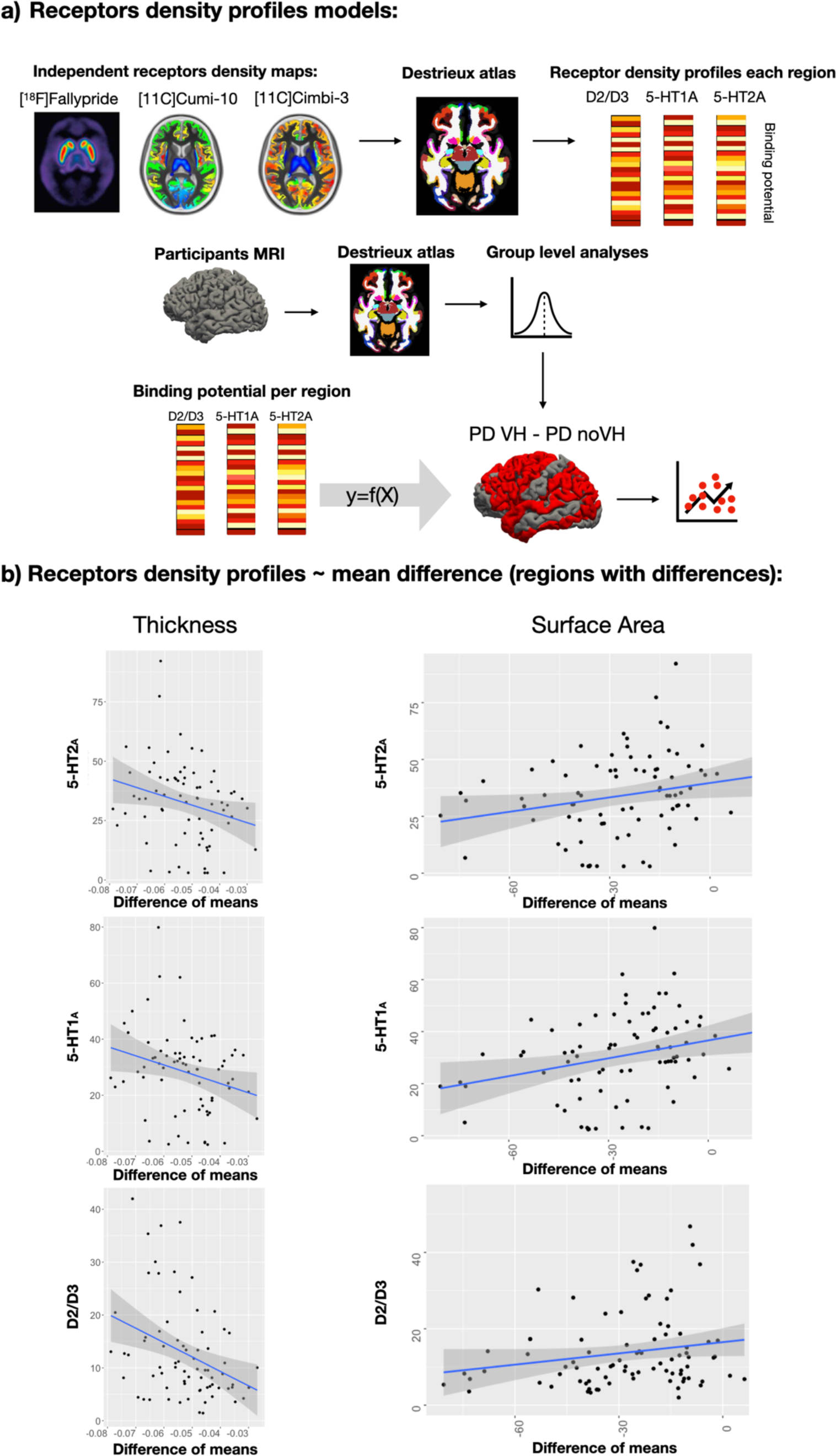
Receptors density profiles: methods and main results. a) Procedure and rationale of the regression models. Both the independent receptor density maps and our participants’ MRI scans were parcellated with the Destrieux atlas. Cortical thickness and SA values were extracted for each region of the atlas for the participants’ scans, and binding potential was extracted for each region of the atlas for the receptor density maps. Each receptor’s binding potential was used in separate models as a predictor of difference of the means of thickness/SA between PD-VH and PD-noVH. b) Results of regression models. Shown are the results of the models with the regions which were different between groups as dependent variable. Results are reported for 5-HT_2A_, 5-HT_1A_ and D2/D3 receptors binding potential and thickness on the left and binding potential and SA on the right.

**Figure 3.**
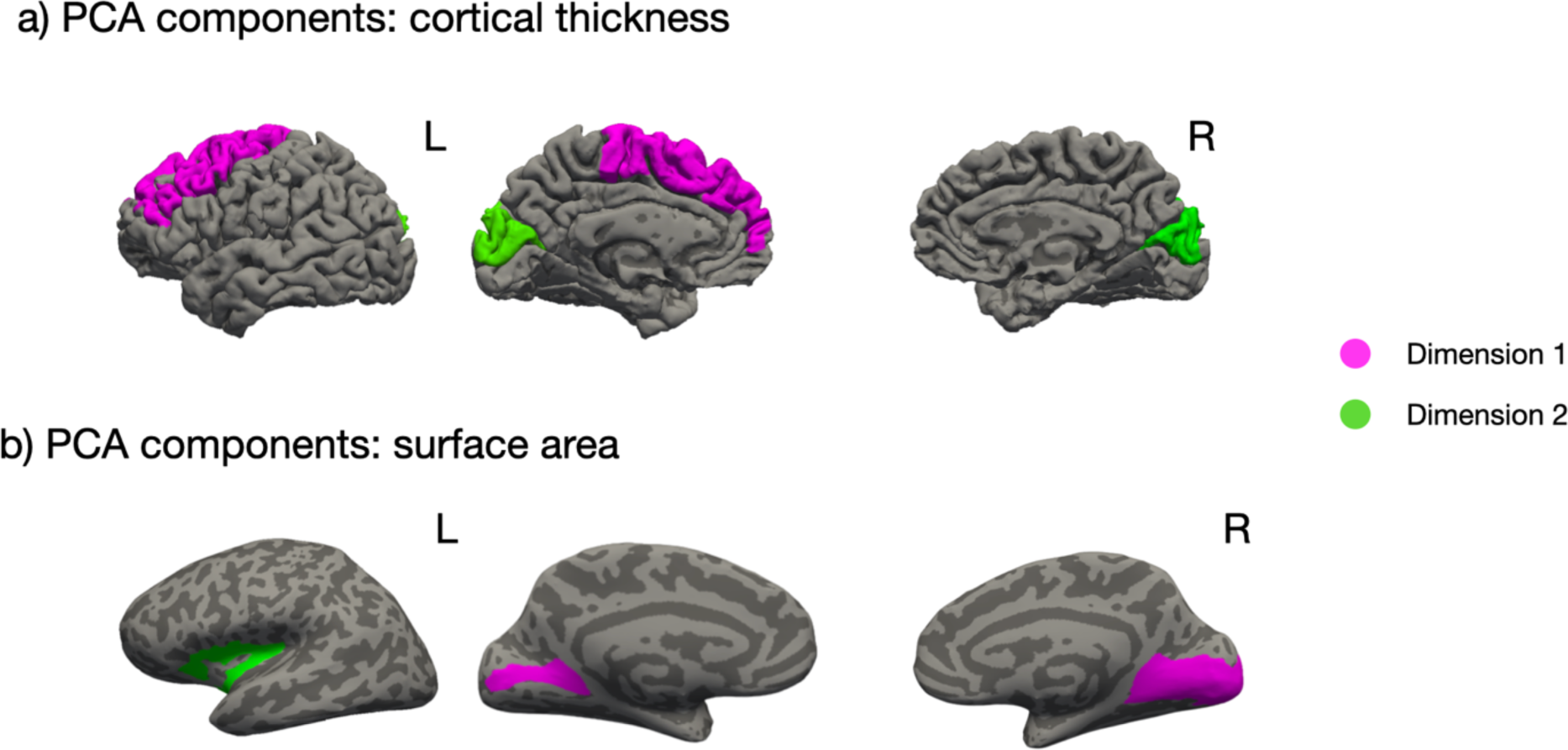
Graphical representation of the regions contribution to each of the Dimensions resulting from the PCA. a) Regions contributing to dimension1 and 2, cortical thickness. Dimension 1 (pink): left superior frontal gyrus, the left middle frontal gyrus and the bilateral precentral gyrus. Dimension 2 (green): bilateral cuneus and occipital superior gyrus. B) Regions contributing to dimension 1 and 2 for surface area. Dimension 1 (pink): left and right calcarine sulci, right occipitotemporal lingual gyrus, right occipital pole, Dimension 2 (green) left central insular area, anterior and superior portions of the circular sulcus of the insula.>

#### Surface area

The models with 5-HT_2A_ binding potential per region as predictor and the mean difference per region as dependent variable was significant, but only for regions which differed between groups (*β* = -.22, *t* = 2.1, *p* =.038). No relationship was observed with all regions included (*β* =.15, t = 1.75, p= .08). The models with 5-HT_1A_ binding potential as predictor was significant for differing regions (*β* =.27, slope= 0.22, *t* = 2.2, *p* =.01) and with lower significance for all regions (*β* =.181, *t* = 2.07, *p* = .04). When using D2/D3 as a predictor, the model was significant for significantly differing regions (*β* =.318, *t* = 2.5, *p* = .01) and the model for all regions showed greater significance (*β* =.277, *t* = 3.24, *p* = .001). In all cases, the greater the mean difference, the lower the binding potential. However, when estimating the confidence intervals of the models, the model for D2/D3 was no longer significant (**S6**). 5-HT_2A_, 5-HT_1A_ and D2D3 slopes for significant regions did not differ (**S6**). See Figure 2, for methods and results and **S6** for further details.

The same models were carried out also for subcortical volumes not yielding significant results for all receptors (see **S6**).

### 2.5 Principal components analysis (PCA)

We performed PCA to reduce the dimensionality of the dataset while preserving variability, to identify underlying clusters to clarify the results from the group-level analyses.

#### Cortical thickness

The PCA returned two dimensions with eigenvalues > 1 explaining 67.58% of total variance. The regions best representing Dimension 1 (eigenvalue = 4.47, 49.69% of variance) as assessed with the cosine squared index were the left superior frontal gyrus, the left middle frontal gyrus and the bilateral precentral gyrus. The regions best representing Dimension 2 (eigenvalue= 1.61, 17.89% of variance) were the cuneus, and the occipital superior gyrus, bilaterally (see **Figure 3a**; for scree plots see **S7**).

#### Surface area

The PCA returned two components with eigenvalues > 1. Dimension 1 (eigenvalue =4.48, 56.01% of variance) and Dimension 2 (eigenvalue = 1.38, 17.21% % of variance) for a total cumulative variance of 73.23% of explained variance. For Dimension 1, the contributing regions were visual regions: left and right calcarine sulci, the right occipitotemporal lingual gyrus and the right occipital pole. For Dimension 2, the contributing regions were the left central insular area, the anterior and superior portions of the circular sulcus of the insula (**Figure 3.b**, **S7**).

For cortical thickness only, we found a significant inverse correlation of Dimension 1 individual contributions and NPI score (*r* = .-138, *p* = .049). In addition, the thickness for the Dimension 1 regions (left SFG, MFG, precentral) negatively correlated with NPI score (*r* =-.15, *p* = .046, one-tailed Pearson correlation), with the individuals having higher pathological score having also the lower thickness in these regions. No significant results were found for surface area, as in the NPI correlational analyses previously reported.

### 2.6 Structural covariance analysis

To explore and characterise the gray matter network-level organisation of PD-VH and PD-noVH patients for cortical thickness and surface area we carried out structural covariance analyses, that assess the covariation of differences in grey matter morphology between different brain structures. After specifying a general linear model for each region, the structural covariance matrices (68x68) of the two groups were defined by estimating the inter-regional correlation between model residuals of thickness and area (in separate models).

#### Cortical thickness

Significant difference of the two covariance matrices (PD-VH, PD-noVH) was first tested (**χ**^2^ =3010.82, *df* = 2278, *z* of differences = 3.83). The cell-by-cell comparisons of residuals’ inter-regional correlation coefficients highlighted differences in the interregional covariance, in particular in the left inferior temporal gyrus, supramarginal gyrus and inferior parietal lobe (IPL), superior frontal (SFG) and inferior frontal gyrus (IFG) pars opercularis and the fusiform and lateral occipital gyri on the right (**Figure 4**). Overall, inter-regional correlations were greater for the PD-VH group (but see also **S8**).

**Figure 4.**
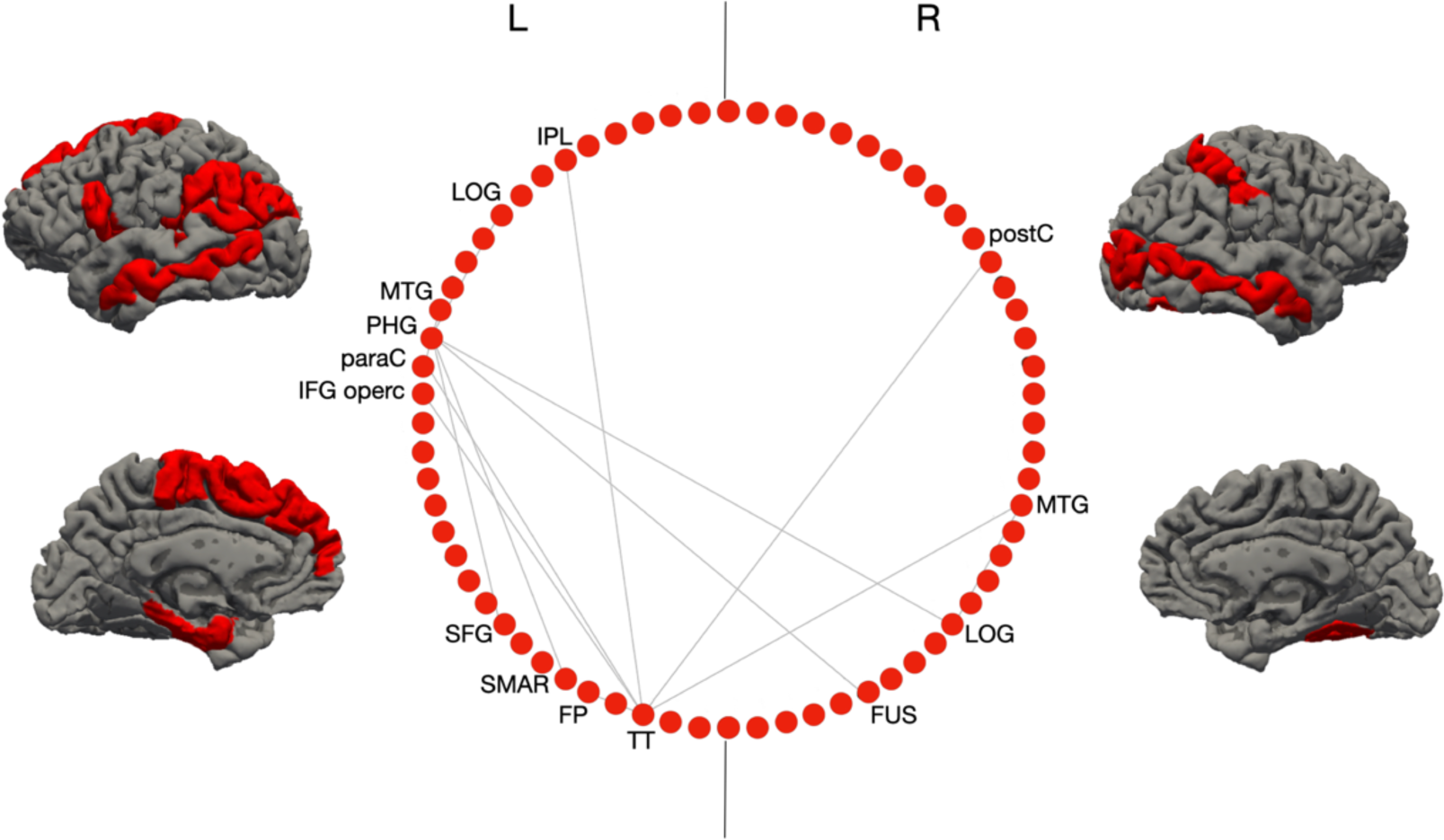
Regions with the most significant difference in inter-regional correlations of cortical thickness between groups: the inter-regional correlations for these regions were significantly greater for VH patients. Shown in the circular plot, only the inter-regional correlations with a difference greater than 0.3 in the r^2^ (for details and z scores on all differences, see **S8**). Legend: IPL = inferior parietal lobule; LOG = lat. Occipital gyrus; MTG = middle temporal gyrus; PHG = parahippocampal gyrus; paraC = paracentral gyrus; IFG opercularis. = inferior frontal gyrus; SFG = superior frontal gyrus; SMAR = supramarginal gyrus; FP = frontoparietal thickness; TT =temporal transverse; FUS = fusiform gyrus; postC = postcentral gyrus. The two vertical lines separate L and R hemisphere regions (left on left).

Hubs, that is nodes (here regions) that are thought to strongly contribute to the global network function, were identified in frontal, parietal and occipital regions for the PD-noVH group, and in frontal, temporal and parietal regions for the PD-VH group (**Figure 5a**). Permutation tests for vertex-level measures returned differences in betweenness centrality, which was greater in PD-VH in the left and right lingual gyrus, in the left lateral occipital gyrus and the right SPL (*p FDR* <.05). Communities are sets of brain regions characterised by denser and stronger relations among themselves, if compared with regions of other communities. Structural covariance-based communities have been found to replicate neighbourhoods observed with seed-based approaches in fMRI and DTI (see Methods for details). The first community in the PD-VH group comprised mainly occipitotemporal regions, with the second involving parietal and some frontal regions. In the PD-noVH group, the first community consisted of mostly frontoparietal regions whereas the second comprised occipitoparietal regions (**Figure 5b**). In addition, the PD-noVH group showed higher modularity, as assessed with bootstrapping (*mean* = 0.29 *SD* = 0.02, *CI* 0.25, 0.36 at density 13%) (for communities by lobe, see **S8**).

**Figure 5.**
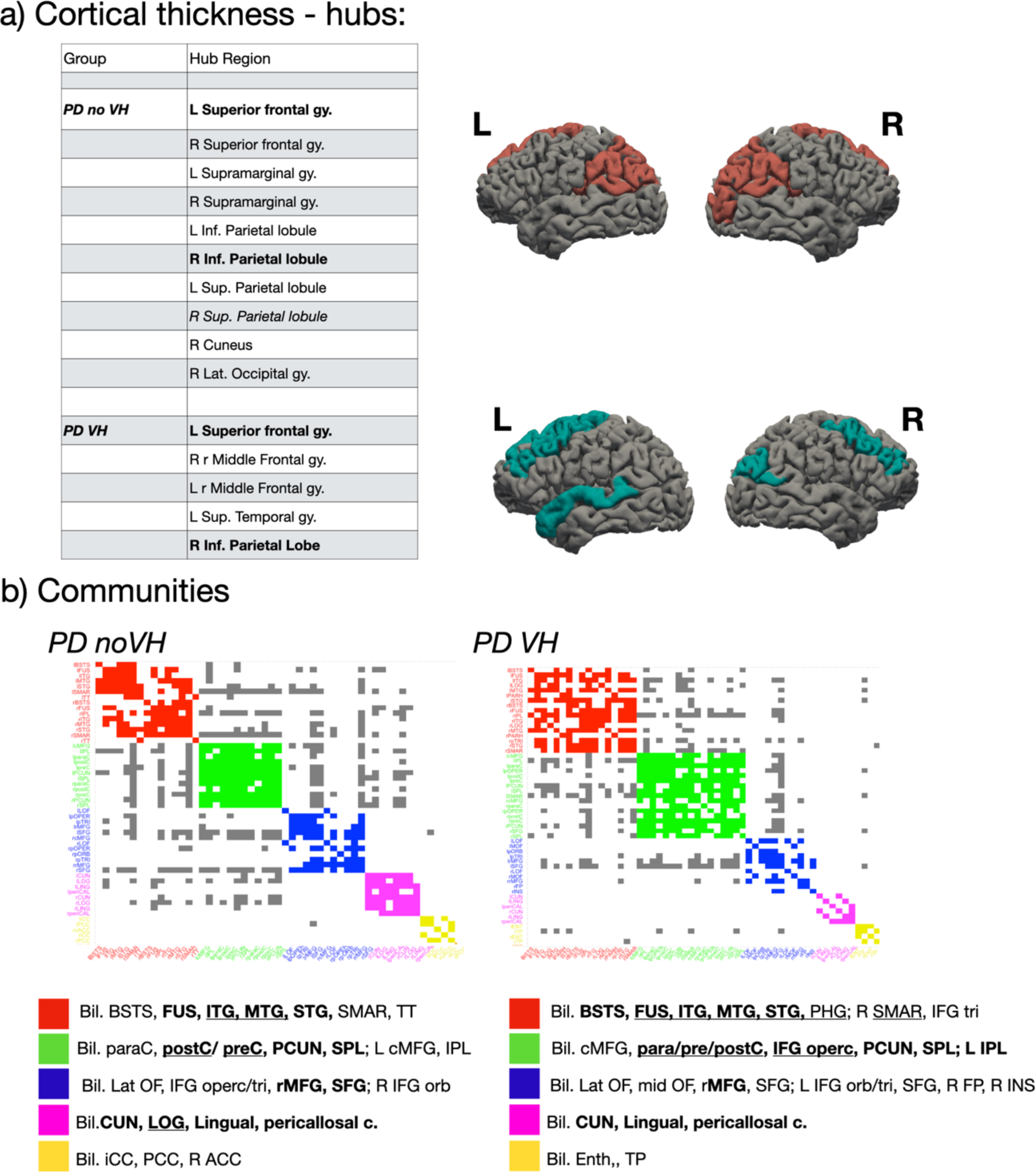
Hubs and communities: cortical thickness. **a)** Hubs identified based on efficiency, betweenness centrality and degree. Regions in **bold** are common hubs for both VH and noVH. **b)** Communities identified for each group. Legend: red = 1^st^ community, green =2nd, blue= 3rd, pink = 4th, yellow = 5th. Only the first five communities are represented as they are the most informative ones. In **bold** the regions identified for that same community also in the surface area analysis. The <u>regions underlined</u> are the same regions presented in the figure with the regions with the greatest difference in inter-regional covariance. Legend: BSTS = banks superior temporal; IPL = inferior parietal lobule; SPL = superior parietal lobule; LOG = lat. Occipital gyrus; MTG = middle temporal gyrus; STG = superior temporal gyrus; cMFG = caudal middle frontal gyrus; PHG = parahippocampal gyrus; paraC = paracentral gyrus; preC = precentral gyrus; postC = postcentral gyrus; IFG = inferior frontal gyrus; SFG = superior frontal gyrus; SMAR = supramarginal gyrus; FP = frontoparietal thickness; TT =temporal transverse; FUS = fusiform gyrus; CUN= cuneus.

Finally, we found no significant correlation between difference of the means in thickness and difference of the means in graph-level measures.

#### Surface area

As for thickness, the two covariance matrices were different (**χ**^2^ = 5347.2, *df* = 2278, *z* of differences = 6.8). In addition, among the others, significant differences in interregional covariance were observed bilaterally in the rostral MFG, STS, fusiform gyrus, and IPL; in the left caudal MFG, lateral occipital gyrus, SPL, and insula and in the right anterior and posterior cingulate, and IFG pars opercularis, with a pattern very similar to the one observed for thickness (see **Figure 6** and **S8**).

**Figure 6.:**
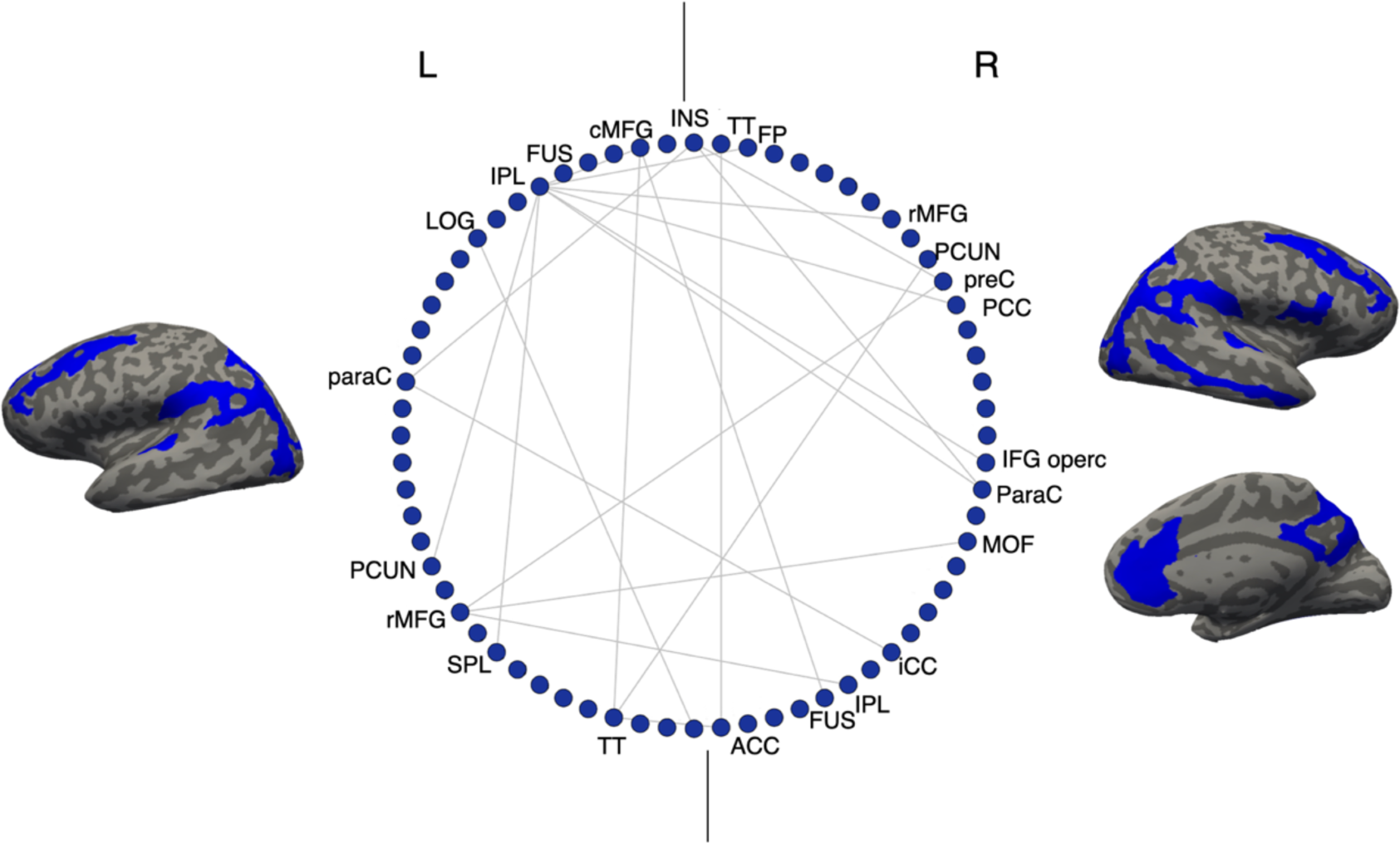
Regions with the most significant difference in inter-regional correlations of surface area between the groups: these correlations were significantly greater for PD-VH. Only the inter-regional correlations with a difference greater than 0.3 in the r^2^ are shown (for details see **S8**). Legend: IPL = inferior parietal lobule; SPL = superior parietal lobule; LOG = lat. Occipital gyrus; MTG = middle temporal gyrus; STG = superior temporal gyrus; cMFG = caudal middle frontal gyrus; PHG = parahippocampal gyrus; paraC = paracentral gyrus; preC = precentral gyrus; IFG = inferior frontal gyrus; SFG = superior frontal gyrus; SMAR = supramarginal gyrus; FP = frontoparietal thickness; TT = temporal transverse; FUS = fusiform gyrus; CUN= cuneus; PCUN = precuneus; MOF = middle orbitofrontal gyrus. The two vertical lines separate L and R hemisphere regions (left on left)

Hubs were identified mainly in occipitotemporal and frontal regions for the PD-noVH group and in frontal, temporal and occipital regions for the PD-VH group, also found in the PCA (**Figure 7**). In accordance with this result, vertex-level permutation tests returned differences in betweenness centrality the left fusiform gyrus; in addition, differences were observed for the middle orbitofrontal gyrus, IFG orbitalis and triangularis, and in the bilateral anterior cingulate (*p* < .003, *p*FDR <.09), whereby centrality was greater for PD-noVH in these regions, but greater for PD-VH in the left caudal MFG and in the right SFG. The first community in the PD-VH group is characterised by occipitotemporal and frontal and the second community by occipito-parietal and parietal regions only (**Figure 7b**; representation by lobe is in S8). In addition, PD-noVH showed greater modularity, as assessed with bootstrapping (0.29, CI 0.21, 0.36 density 13%).

**Figure 7.**
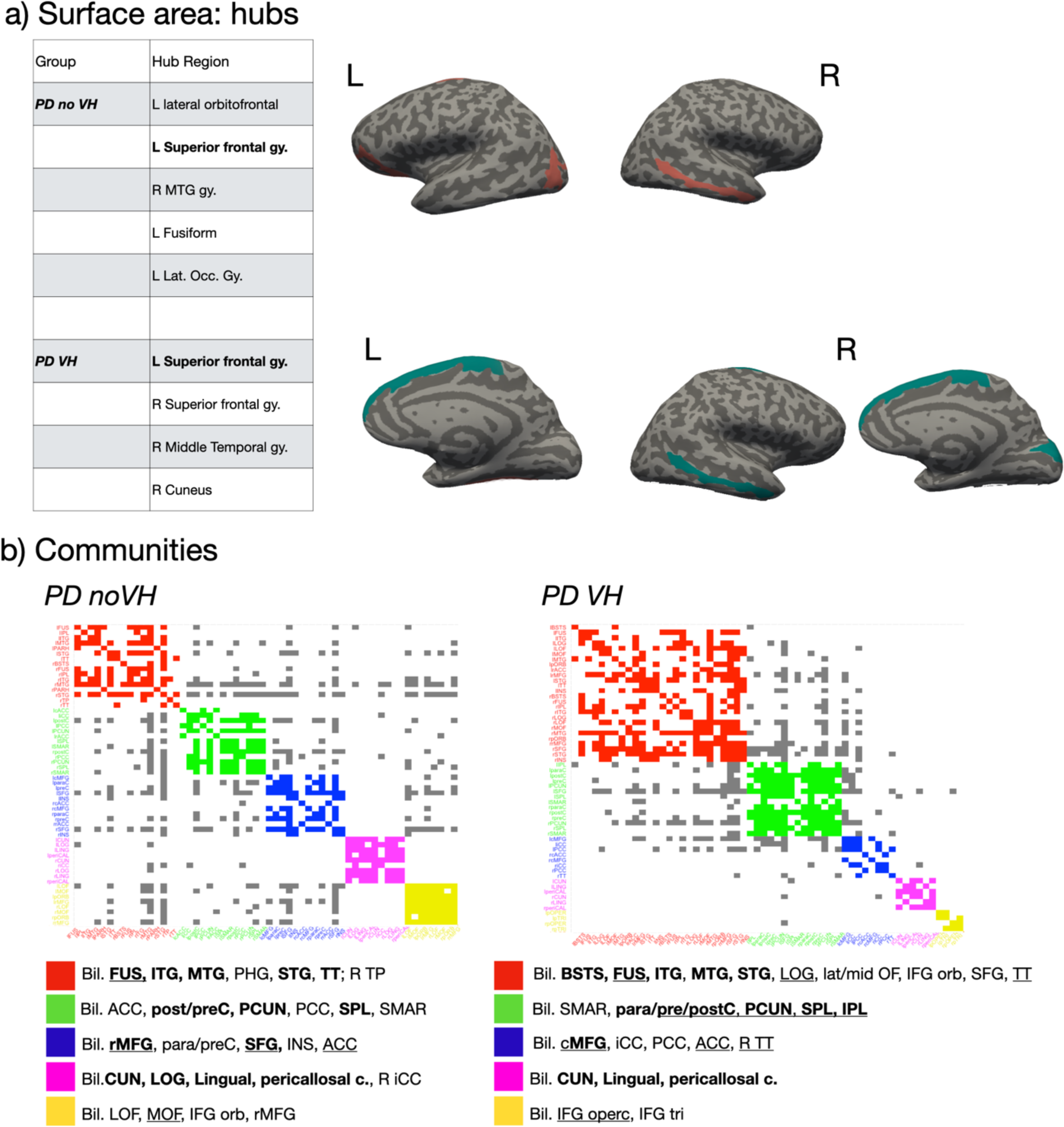
Hubs and communities: surface area. **a)** SA hubs identified based on efficiency, betweenness centrality and degree. Regions in **bold** are common hubs for both VH and noVH patients. **b)** Communities identified for each group. Legend: red = 1^st^ community, green = 2nd, blue= 3rd, pink = 4th, yellow = 5^th^. In **bold** the regions identified for that same community also in the SA analysis. The <u>regions underlined</u> are the same regions presented in the figure with the regions with the greatest difference in inter-regional covariance. Legend: BSTS = banks superior temporal; IPL = inferior parietal lobule; SPL = superior parietal lobule; LOG = lat. Occipital gyrus; MTG = middle temporal gyrus; STG = superior temporal gyrus; ITG = inferior temporal gyrus; cMFG = caudal middle frontal gyrus; PHG = parahippocampal gyrus; paraC = paracentral gyrus; preC = precentral gyrus; postC = postcentral gyrus; IFG = inferior frontal gyrus; SFG = superior frontal gyrus; SMAR = supramarginal gyrus; FP = frontoparietal thickness; TT = temporal transverse; FUS = fusiform gyrus; CUN= cuneus

Finally, we found a significant positive correlation between difference of the means in the NPI subsample and with the difference in local efficiency (*r* = .24, *p* = 0.02), whereby the greater the difference in the surface area, the greater the difference in the local efficiency. The regions with both the greatest area differences and efficiency differences were the bilateral lingual gyrus, lateral occipital gyrus, right cuneus and right insula.

## 3. Discussion

We have presented a mega-analysis of patients with Parkinson’s disease with and without visual hallucinations, demonstrating widespread alterations in brain structure, with differential effects for cortical thickness and surface area and examined their relationship to receptor distributions and network-level effects. Below we discuss the implications of the findings and their relationship to current theories of VH.

### Cortical thickness and surface area

Cortical thickness and surface area (SA) are considered two separate components in ageing and disease (Dickerson et al. 2009; Storsve et al. 2014; Cox et al., 2018) reflecting different aspects of the neurodegenerative process. Thickness loss relates to cortical layering and, by inference, cytoarchitecture, while surface area relates to gyral anatomy and, by inference, underlying white matter. Widespread reductions in *cortical thickness* in hallucinators were identified in the occipital, parietal, temporal, frontal and limbic lobes. The regions of reduced thickness encompassed all cortical regions identified in previous structural imaging studies (for a review, Lenka et al., 2015), suggesting previous variability may relate to stochastic effects introduced by relatively smaller samples and design differences. With the larger sample of the mega-analysis, the extent of cortical regions involved appears wider than previously suspected. However, not all regions are equally affected and, notably, there appears to be a posterior asymmetry with relative sparing of the left ventral visual stream (ventral occipito-temporal cortex) compared to the homologous region in the right hemisphere. This region plays a key role in all models of VH in PD but a greater involvement of the right hemisphere has not been noted previously. The PCA analysis helped define key sub-regions within the extensive areas of cortical thinning that contributed most to the group difference, identifying a frontal and an occipital dimension. Of these, the cuneus bilaterally and left dorso-medial aspect of the superior frontal gyrus emerged as the dominant components. These regions have been reported in previous studies but do not play a prominent role in accounts of VH in PD. The cuneus is one of the earliest regions to show cortical atrophy in PDP, present at the earliest stages when only minor hallucinations occur (Pagonabarraga et al., 2014), while cortical thinning in the dorso-medial superior frontal gyrus has been reported in patients, months to years prior to the development of VH (ffytche et al., 2017). It may be that the prominence of these regions in the mega-analysis relates to the longer duration of these changes compared to other brain regions resulting in a greater consistency of thickness reduction between patients.

For *SA,* the differences between groups were more circumscribed with bilateral medial occipital SA reduction for patients with VH in a region corresponding to the primary visual cortex and its surrounds (striate and extra-striate cortex) and the left insula. This is the first-time such extensive structural changes have been identified in the primary visual cortex and its surrounds in PD patients with VH and helps account for wide-ranging low-level visual deficits found (see Weil et al., 2016 for a review). These regions also have reduced cortical thickness but their prominence in the SA analysis may imply additional gyral atrophy, sulcal widening and a reduction of underlying white matter.

The mega-analysis also allowed us to move beyond a binary comparison of VH versus noVH to examine brain regions linked to VH severity as measured by the NPI hallucination subscale score (a composite score derived from the product of frequency and distress ratings) and taking into account any variability associated to age, gender, TIV, medication, cognition, disease onset and PD severity. Regions with reduced thickness for higher severity scores (negative correlation) were found in posterior parietal, posterior cingulate and superior temporal cortex while a region in the frontal lobe was found with greater thickness for higher severity scores. Previous studies have associated these regions with mental rotation and visuospatial transformation (Papadopoulus et al., 2018) and imagery (Tian et al., 2016) for the IPS, and biological motion detection (Sokolov et al., 2018) for the STS. These processes are altered in patients with PD and VH (Firbank et al., 2018; Shine et al., 2015), thus one can infer an involvement of these processes and these regions in VH severity. In addition, the IPS is also part of the dorsal attentional network, previously implicated in VH in PD (Shine et al., 2014a; see discussion below). These regions were also identified as hubs in the structural covariance analysis, discussed further below. As separate distress and frequency scores were available only for a part of the subsample, we were unable to analyse the two components of the severity score separately, thus we cannot disentangle whether these correlations are driven primarily by the frequency or distress measures. However, this is the first time a link between cortical structural changes and phenomenological aspects of VH severity has been identified.

### Subcortical regions, hippocampus and cerebellum

In addition to the detailed analysis of the cerebral cortex we were able to examine the volumes of subcortical structures as well. Bilateral volume reduction was found in the amygdalae. Lewy bodies have been found in the basolateral nucleus of the amygdala associated with VH in PD patients at a similar level of cognitive impairment to those studied here (Harding et al., 2002) that may account for this finding. Unlike the amygdala, there are only sparse Lewy bodies in the hippocampus at this disease stage and volume changes in this structure are more difficult to interpret. Since the prevalence of VH increases as PD progresses, tracking cognitive progression from PD-MCI to PD-dementia, it is difficult to disentangle brain changes related primarily to cognitive decline from those related primarily to VH or that may contribute equally to both. Reductions of hippocampal volume (particularly its anterior portions) have been found in some, but not all, studies of VH in PD, depending on whether patients are matched for cognitive decline (e.g. Yao et al., 2016; Ibarrexete-Bilbao et al., 2008). Here we found smaller left (and a trend for right) hippocampus in the NPI sample where we were able to covary for age, gender, TIV, onset, LED, PD severity and cognition. We did not find hippocampal volume reduction in the full data set covarying only for age, gender and TIV. The volume reductions in the NPI analysis cannot be explained by differences in cognition or PD progression between groups, confirming a role for the hippocampus in the mechanism of VH that is independent of cognition (e.g. Ibarrexete-Bilbao et al., 2008), thus highlighting the need to carefully design studies and control for cognitive and disease factors when examining hippocampal contributions to VH. The thalamus has been suggested as a key hub linking several cortical networks associated with VH in PD (Weil et al., 2019). We did not find altered thalamic volumes in PD-VH in the main analyses or subgroup NPI analysis which included a wider range of covariates (S4). This does not rule out involvement of the thalamus in the pathophysiology of VH in PD but does suggest any functional changes in this structure are not associated with volume loss. Finally, reduced volume in cerebellar lobules VIII, IX/VII and Crus 1 is associated with VH in PD (Lawn and ffytche, 2020). Freesurfer does not segment specific cerebellar subfields but volume changes were found in cerebellar white matter that may relate to these cerebellar cortical changes (Lawn and ffytche, 2020).

### Neurotransmitter receptor density and structural imaging changes

There is only sparse Lewy body pathology in the cortex of PD patients with VH at the disease stage included in our analysis (Harding et al., 2002), raising the question of what causes the extensive cortical changes found in this and previous studies. One possibility is that such cortical changes represent synaptic loss secondary to degeneration in neurotransmitter inputs to the cortex. Previous studies have found changes in cholinergic, serotonergic, dopaminergic and GABAergic systems in PD patients with VH (e.g. Firbank et al., 2019; Yasue et al., 2016; Ballanger et al., 2010); however, the relationship between regions of cortex with volume loss and the cortical distribution of these neurotransmitter systems has yet to be examined. We were able to investigate this relationship for subtypes of dopamine and serotonin receptors for which high resolution maps are available and found that cortical regions with higher binding had increased cortical volume loss. The association, in particular for 5-HT_2A_, was confined to regions linked to VH rather than the cortex as a whole, suggesting the neurotransmitter effects were specific to VH, consistent with the possibility that degeneration in these neurotransmitter systems in PD underlies synaptic loss and cortical thinning. Receptor binding maps for D2/D3 and 5-HT_2A_ were not correlated suggesting different cortical regions contributed to the associations found for D2/D3 and 5-HT_2A_. 5-HT_2A_ and 5-HT_1A_ binding maps were correlated so the same cortical regions are likely to have contributed to both serotonin findings. While increased binding was associated with increased thickness loss, the opposite association was found for SA, with higher binding exhibiting less SA reduction. This finding was not specific to VH regions for dopamine and 5-HT_1A_ so may reflect a different process to the thickness alterations found. It is also unclear what causes low binding regions to be associated with increased loss of SA. Finally, there was no suggestion of a greater contribution of one neurotransmitter system over the other to cortical thickness loss, with equivalent slopes for all three receptors maps examined.

### Structural covariance

The examination of inter-regional correlations, with areas sharing reductions in thickness or SA considered part of a functionally connected network, showed that regions of greater inter-regional thickness correlation in PD-VH overlap with those of the dorsal and ventral attention networks (DAN and VAN) (Corbetta et al., 2002), with the notable addition of para-hippocampal regions. Most of these regions of higher covariance have reduced thickness in PD-VH, suggesting the covariance is driven by correlated reductions in thickness. Dysregulation of VAN, DAN and default mode networks (DMN) have been implicated in models of VH in PD (Shine et al., 2014a) with reduced activity in the DAN of PD-VH (Shine et al., 2014b), and the inter-regional covariance findings support this view. In contrast, the inter-regional SA covariance findings highlight key DMN regions in medial frontal and posterior cingulate cortex. These regions were not found to have reduced SA in PD-VH, suggesting a relative preservation of the DMN compared to VAN and DAN. Indeed, results from dynamic fMRI have indicated active coupling between the DMN and the visual network, which correlated with the frequency of misperceptions, as opposed to reduced connectivity between the DMN, VAN and DAN (Shine et al. 2015). Other metrics derived from the covariance structure include hubs defined by the richness of their interconnections and communities defined by their local strength of covariance. Hub metrics for thickness in the occipital lobe and parietal lobe were stronger in patients with VH, suggesting cortical thinning has a wider impact on the network in these patients, highlighting the importance of functional alterations in early visual areas in VH. One could argue that VH may not only depend upon on areas presenting neural pathology, but also on areas that may be relatively unaffected but operate in a network where there is pathology elsewhere, thus becoming functionally pathological while structurally intact (ffytche, 2008). Indeed, all the regions where richness of connections was either lower or higher for PD-VH fell outside areas of reduced SA in VH, suggestive of a more functional pathology which needs to be further explored with functional connectivity. Finally, there were qualitative differences in the communities of highly associated regions for PD-VH compared to PD- noVH in both the thickness and SA analysis. Of particular note was the extent of interconnected areas in the ventral, lateral and medial temporal lobe that was larger in the PD-VH group. These regions had reduced thickness in PD-VH implying the local extent of thickness reduction is greater in PD-VH.

### Strengths and limitations

This is the first mega-analysis of VH in PD, pooling data to create the largest sample of PD patients with and without VH tested to date. While this is a major strength of the study, it also introduces complexities that smaller studies do not have to address. One is the variability of clinical data available for each site, limiting the analyses we could perform with the full dataset of 493 participants. This means that some of the key analyses, for example those related to cognitive covariates and disease duration or symptom scores, could only be carried out in a smaller sample of 146 participants, but this is still substantially larger than any previous study. Another complexity is the need to address variance in the data caused by scanning at different sites and scanner types. Previous studies have typically used voxel-based methods to examine structural differences between PD-VH and PD-noVH. We used a different method to allow us to harmonise data between sites and examine cortical thickness and SA separately, but this means our findings are not directly comparable to those of previous studies. The primary focus of the study is on the cerebral cortex so we have not attempted to examine the detailed anatomy of regions such as the basal ganglia, hippocampus, cerebellum and thalamus that may have a role in VH. Finally, we do not have access to high resolution density maps for cholinergic receptor subtypes which limits the range of neurotransmitter analyses we can perform.

### Conclusions

The mega-analysis has allowed us to resolve several uncertainties in the previous literature and describe new features of the VH phenotype in PD. With a sufficiently large sample, more widely distributed cortical involvement emerges than previously suspected with the important novel finding of involvement of the primary visual cortex and its surrounds. Structural covariance modelling has helped dissect out networks linked to attentional control within the widespread cortical regions affected, adding further evidence for the role of these networks in PD-VH. The findings also help resolve ambiguities between structural correlates of general cognitive decline or PD progression and those specifically related to VH. Patients at the same stage of PD and general cognitive impairment who experience VH have lower hippocampal volumes than those who do not. The hippocampus does not currently play a central role in models of VH in PD and our findings suggest this needs to be reconsidered. We can argue that the hippocampus represents part of an extended DMN composed of functional hubs, a dorsal medial subsystem and a medial temporal subsystem, which includes the hippocampus (Qi et al., 2018). Thus, structural covariance, graph-level analyses and structural hippocampal imaging point to the involvement of attentional control networks in PD-VH. Finally, the findings shed light on why widespread cortical changes occur at a stage of PD with only sparse cortical neuropathology. The associations between dopaminergic and serotonergic receptor binding and cortical thickness provide the first evidence that the cortical changes may be driven by neurotransmitter reductions, raising the possibility of novel interventions to mitigate these effects at an earlier stage of disease.

## Author Contributions

Conceptualisation, MV, DF and MM; Methodology, MV and MM; Investigation, MM, MV, DF; Formal analysis, MV; Writing – Original Draft, MV, MM, DF.; Writing – Review & Editing, PHL, SJC, SL, RW, LG, MH, CM, GM, HM, DP, RJ, KD, JPT, MF, SLH; Visualisation, MV; Data Curation, MV; Funding Acquisition, MV was supported by MRC grant MR/0059311; Resources, PHL, SJC, SL, RW, LG, MH, CM, GM, HM, SLH, DP, RJ, KD, JPT, MF, DF; Supervision, MM.

## Inclusion and Diversity Statement

One or more of the authors of this paper self-identifies as an underrepresented ethnic minority in science. Some of the original studies worked to ensure sex balance in the selection of non-human subjects.

## 4. Methods

*Lead contact:* Further information and request for resources should be directed to and will be fulfilled by the lead author Miriam Vignando.

*Materials availability:* This study did not generate new unique reagents

*Data and code availability:* The datasets generated during this study will be made available upon publication at the project page on the Open Science Framework https://osf.io/fv2k7/registrations.

### 4.1 Studies selection

Based on the literature, we identified N=17 studies of VH in patients with PD that included acquisition of T1-weighted structural MRI scan, as part of a structural or functional data analyses, and with patients meeting our inclusion criteria (see below). We contacted the research groups responsible for the studies and among those N=8 groups took part in the project, offering previously published and/or unpublished data: Prof. Simon Lewis (University of Sydney, Shine et al. 2014 and unpublished data), Prof. Phil Hyu Lee and Dr. Chung (Yonsei University, Shin et al., 2012), Prof. Henry Mak, Prof. Grainne McAlonan and Prof. S.L. Ho (King’s College London and The University of Hong Kong, Yao et al., 2014), Prof. Kathy Dujardin, Prof. Renaud Jardri and Dr. Delphine Pins (University of Lille, Lefebvre et al., 2016), Prof. John-Paul Taylor and Dr. Michael Firbank (Newcastle University, Firbank et al., 2018), Dr. Rimona Weil (University College London, sample in Zarkali et al., 2020, T1-weighted data submitted), Prof. Michele Hu, Prof. Clare Mackay and Dr. Ludovica Griffanti (Oxford Parkinson’s Centre Discovery Cohort, Baig et al., 2015; Griffanti et al., 2020), Dr. Dominic ffytche (King’s College London, unpublished data from PDHAL study) (see Table 1 in the Results section for details). Only data from participants diagnosed as dementia-free were included to minimise the contribution to the study of global cortical changes in patients with PD dementia. Ethical approval for the study (LRS-19/20-17680) was given on the 25/03/2020 by King’s College London Research Ethics Office, Psychiatry, Nursing and Midwifery (PNM) Research Ethics Panel. The study was subsequently pre-registered on the Open Science Framework site on 04/05/2020 (https://osf.io/nzatk). The methods follow the pre-registered plan with the addition of exploratory graph theoretical analyses based on structural covariance (section 4.3.4 and results in section 2.6).

### 4.2 Participants

Raw T1-weighted MRI scans were obtained from 8 different groups for a total of 519 subjects. We used 493 MRI scans in the analysis after discarding N=20 participants who did not meet the criteria in terms of diagnosis (e.g. healthy controls, with diagnosis of dementia) or whose scan did not segment well during pre-processing and subsequent troubleshooting steps or was not suitable for analysis (e.g. motion) (N=6). Patients with a MMSE score below 24 (raw) were retained (N=8) only when part of a published work in which the absence of dementia was specifically stated. The final sample comprised 493 participants, 135 with VH, 358 without VH (further details in *Results* section and in **Table 1** and **S2**). Hallucination data collection varied across groups, as several used a different scale to screen for VH. Each group had previously divided patients into PD-VH and PD-noVH and we retained these original groupings for the mega-analysis.

### 4.3 MRI data pre-processing and harmonisation

MRI data was pre-processed with Freesurfer 6.0.0 (Fischl, 2012; Dale et al., 1999) to estimate cortical thickness, surface area and subcortical volumes. Data was processed on King’s College London HPC infrastructure Rosalind (https://rosalind.kcl.ac.uk), with the standard recon-all procedure, consisting of motion correction, skull-stripping, affine registration to Talairach atlas, segmentation, smoothing, and parcellation mapping. In order to screen for possible errors in the segmentation process, mean cortical thickness measures and manual slice by slice inspection were used to identify possible errors in the white-grey matter boundary and pial reconstruction steps. For subjects that did not segment properly the failed processing steps were re-run (autorecon3) after performing the appropriate corrections. Low quality scans (e.g. with excessive motion, n= 4) or scans that did not segment well upon troubleshooting (n =2) were discarded. Individual cortical thickness, subcortical volumes and surface area measures were extracted based on the Destrieux atlas (Destrieux et al., 2010). In order to explore structural differences between patients with and without VH across the different cohorts minimising variance due to different recruitment sites and, therefore, different scanners, we used a harmonisation method. ComBat is an empirical Bayesian algorithm aiming at minimising the variance due to the scanner features and to maintain the variance related to biological features and has been previously successfully used in studies of cortical thickness (Fortin et al., 2018; Radua et al., 2020). In this study, this method has been also used to harmonise volume and surface area for each participant (see **Supplemental information S1** for more details about this method and plotted results).

#### 4.3.1 Group differences analysis

First, we conducted a meta-analysis with R package ‘metafor’ (Vieachtbauer, 2010) to check whether patients were matched on the relevant demographic and clinical variables. Results are mentioned in the main text and reported with forest plots and a detailed description in **Supplemental Information S2**.

Then, we conducted separate exploratory ANOVAs and MANCOVAs for cortical thickness, surface area and subcortical volumes to screen for group differences between hallucinators and non-hallucinators, with age, gender and total intracranial volume (TIV) as covariates (when appropriate upon checking assumptions; see *Results* and Supplemental Information **S2**). Multiple comparisons were Bonferroni corrected. The models were calculated using SPSS 24.0.0.0 (IBM corp. 2016) and R 4.0.0 (R core team, 2017). Results are presented in **Figure 1**, created with a custom colour coding based on *p* values and by overlaying region labels on a brain render.

We used Tukey’s method programmed in R with the 1.5*IQR rule to identify outliers other than those excluded upon unsuccessful pre-processing. This allowed the careful inspection of the identified subjects in order to verify whether the outlier value depended upon measure errors (e.g. harmonisation bugs) or incorrectly entered data, or on the subject, with the purpose of retaining outliers depending on the subject (e.g. intrinsic features of the subject). No participants were discarded upon this check for this analysis.

#### 4.3.2 Sensitivity and Subgroup analysis

Of the eight original groups, three used the Neuropsychiatric Inventory (NPI) to score visual hallucinations. For this subgroup of studies, patients were matched for age, gender, onset, levodopa equivalent daily dose (LEDD), and Mini Mental State Examination (MMSE) score. Within each of the 3 studies, patients were also matched in terms of motor symptoms severity (UPDRS-III). We also ran a one-way ANOVA to check whether the subsample was matched for UPDRS-III but data was missing for 20 participants. We computed the group mean and used that to fill the missing value for the between groups multivariate ANOVA. We carried out Pearson’s product moment correlation coefficient between NPI score and the cortical thickness, surface area and subcortical volume data; we computed the same analysis with LED as a covariate in order to address its potential role in VH severity. In addition, we compared the PD-VH and PD-noVH in the data set using the original VH binary scores to check for consistency in the results with the larger data set, including age, gender, disease onset, LED, PD severity (UPDRS-III) and MMSE as covariates (**Supplemental Information S4**). We also conducted analyses of variance with a larger subgroup and with graded VH scores (mild, moderate, severe), together with an ordinal logistic regression (for details on the sample, methods and results see **Supplemental Information S5**).

#### 4.3.3 Receptor density profiles

Regression models with the difference of the means (VH – noVH) of morphometrical features (thickness, surface area, subcortical volume) as dependent variable and receptors density profiles as predictors were carried out, with a methodology similar to (Selvaggi et al., 2018). Specifically, receptors density profiles were obtained for D2/D3, 5-HT_1A_ and 5-HT_2A_ based on a [^18^F] Fallypride template (Dunn et al., 2013) and a [11C] Cumi-101 5-HT_1A_ and a [11C] Cimbi-36 5-HT_2A_ templates (Beliveau et al., 2017), respectively. We have focussed on DA and 5-HT as high resolution templates are available for these receptors of interest at the moment. Including cholinergic maps in the analysis would greatly enrich this approach given the importance of cholinergic transmission in VH in PD (Perez-Lloret and Barrantes, 2016), and will be done once high resolutions templates will be available. [^18^F] Fallypride is a D2/D3 receptor antagonist with a high signal to noise ratio (Mukherjee et al., 2002). [11C] Cumi-101 and [11C] Cimbi-36 are high affinity PET radioligands for 5-HT_1A_ and 5-HT_2A_ receptors (Beliveau et al., 2017). Parametric modelling of the binding potential used the cerebellum as reference region (Ichise et al., 2003) and thus the vertices corresponding to the cerebellum were excluded from the regression analyses. Each of these templates was registered to the Talairach space using the *fsaverage* template subject and parcellated with the Destrieux atlas, to ensure alignment with the parcellated structural data of our participants. For each of the vertices we extracted the binding potential using *fslmeants*. Regression models were carried out to estimate the relationship between cortical thickness and surface area differences of the mean between VH and noVH patients (regions resulting from the first group-level MANCOVAs and ANOVAs, see **2.1**, **S3**) and receptor density profiles. For surface area, we used regions that resulted different in PD-VH vs. PD-noVH from an exploratory one-way ANOVA, as the number of regions resulting different in the basis of the MANCOVAs performed and reported in S3 were too small in number to carry out a more powered model. We ran separate models for each receptor and for thickness, surface area and volume. In addition, for each receptor we ran three different models. First, we examined the relationship between the receptor’s binding potential in the regions with significant differences in cortical thickness/surface area between PD-VH and PD-noVH. The sloopes for these models were also compared (**Supplemental Information S6**). Then, in order to better investigate such relationship, we also assessed whether the receptor’s binding potential could predict thickness/area values for all regions; finally, with the same purpose, we ran models considering only regions where the difference between the groups was not significant (**S6**). Linear regression models were coded in R using the packages rstatix (Kassambara, 2020) and MASS (Venables, W. N. & Ripley, B. D. 2002). For each regression model, in order to identify outliers, Cook’s distance was computed and any data point with a Cook’s distance >1 was marked as highly influential, explored and if appropriate discarded (Cook and Weisberg, 1982). In addition, the confidence intervals of the significant regression models were estimated with the bootstrapping technique (Efron and Tibishirani, 1986) with 100,000 cycles (**S6**). Methods and results for thickness and surface area are graphically represented in Figure 3, for results on volume and further details see Supplemental Information S6).

#### 4.3.4 Principal component analysis (PCA)

Results from the MANCOVAs comparing PD-VH and PD-noVH highlighted the involvement of widespread cortical regions in a high dimensional dataset. We used principal component analysis (PCA), in order to reduce the dimensionality of the dataset and to identify putative latent dimensions underlying the differences in structure in PD-VH versus PD-noVH patients while retaining as much variance as possible (Joliffe and Cadima, 2016). Data from both hemispheres was entered in each model (one for cortical thickness, one for surface area). Analyses were carried out with R packages *factominer* (Le, Josse, Husson, 2008) and *factoextra* (Kassambara and Mundt, 2017). The scree plots for the PCA are reported in **S7**. Separate PCAs were carried out for thickness and surface area. PCA inputs comprised the significantly different regions from the MANCOVAs (**S3**). Results are presented in Figure 3, created with a custom colour coding based on the components and by overlaying region labels on a brain render. To further explore a possible relationship of PCA components and hallucination severity, we carried out correlational analyses (Pearson product-moment) between the individual contributions to the different PCA dimensions, the NPI scores in the NPI subsample and the mean thickness and surface area of each dimension/component, that is the mean thickness/area across the regions constituting that component.

#### 4.3.5 Structural covariance and graph theory analysis

In order to investigate inter-regional properties to explore and characterise the gray matter network-level organisation of PD-VH, we built networks based on structural covariance, a technique that assays covariation of differences in grey matter morphology between different brain structures across a specific population (Lerch et al., 2017; 2006). Since the most widely used atlas for this kind of analysis is the Desikan-Killiany atlas (Desikan et al., 2006; see Carmon et al., 2020), we extracted morphometric features (thickness, surface area) at the 68 vertices of this atlas. The dataset was harmonised for multi-site effects with the same procedure described in section *4.3.1*. The dataset was reduced to 467 cases as the design matrix based on the full dataset was not invertible due to high collinearity of some columns. We discarded N=26 subjects comingfrom the smallest datasets and the problem was overridden. Homogeneity of groups was verified with a Levene’s test (Nimon, 2012; Cheung, 2019).

The dataset counts 467 subjects, 118 PD-VH, 349 PD-noVH, with participants being matched for age. *Age* and *gender* were used as covariates in the models. Analyses were carried out with R package *braingraph* (Watson, 2018; Watson et al., 2018) and *igraph* (Csardi and Nepuz, 2006). To construct the networks, first we specified a general linear model for each region (thickness/area as outcome variable, age and gender as covariates). The structural covariance matrices of the two groups were defined by estimating the inter-regional correlation between model residuals of thickness and area (in separate models) (e.g. He et al., 2007) to build a 68x68 matrix. A density-based threshold (Fornito, Zalesky and Bullmore, 2016) was applied to the matrix in order to retain a percentage of the most positive correlations as non-zero elements in a binary adjacency matrix. Different densities ranging from 0.05 to 0.20 with a 0.01 step size were explored. The differences between PD-VH and PD-noVH covariance matrices were then computed, first to establish that the two matrices differed significantly from one another; secondly, a cell by cell comparison was carried out to establish which covariance patterns were significantly greater for the PD-VH group compared to the PD-noVH group. Random undirected and unweighted graphs were created for each group, and vertex- and graph-level metrics were computed for each group. For visualisation purposes a density of 0.13 was selected. Vertex importance was assessed using degree, betweenness centrality and nodal efficiency. A hub was categorised as such if its betweenness centrality was greater than the mean plus 1 standard deviation - calculated on all vertices at the same density (e.g. Bernhardt et al., 2011; Hosseini et al., 2013; Tijms et al., 2013; Wang et al., 2013; Watson et al., 2018). To assess network segregation in order to better understand the communities observed, we used modularity, which is a measure of the strength of network partitions. High modularity is a measure of how much vertices from the same community are more connected to each other. Modularity was computed with the Louvain algorithms, which also partitioned the network in communities (Blondel, Guillaume, Lambiotte and Lefebvre, 2008). Cortical thickness-based networks have been shown to have distinct modules/communities of regions, similar to those derived from fMRI and DTI data (see Watson et al., 2018). Network analyses were performed with permutation tests (5000 cycles) and bootstrapping analyses to compare vertex-level measures. Results were false discovery rate corrected.

Finally, to further assess the relationship between graph level metrics and visual hallucinations in the full sample, we computed Pearson’s correlation coefficients between the difference of the means of graph metrics of interest (vulnerability, transitivity, local and nodal efficiency, path length, betweenness centrality, eccentricity, distance) for the models on thickness and surface area separately, and the difference of the means in thickness and in surface area, respectively, with the NPI subsample, for which we have all clinical and demographic information and in which participants are matched on all those variables.

## Data Availability

Datasets generated during this study will be made available upon publication at the project page on the Open Science Framework https://osf.io/fv2k7/registrations.

## Acknowledgments

Dr. Miriam Vignando was supported by the MRC grant MR/0059311. Prof. John-Paul Taylor and Dr. Firbank’s study was supported by the Northumberland Tyne and Wear NHS Foundation Trust, and by the National Institute for Health Research (NIHR) Newcastle Biomedical Research Centre (BRC) in Newcastle upon Tyne Hospitals NHS Foundation Trust and Newcastle University. Prof. Simon Lewis wishes to thank the NHMRC for the Dementia Research Team Grant, NHMRC Project Grant, Parkinson’s NSW Seed Grant 2013, NHMRC Investigator grant, NHMRC-ARC Dementia Fellowship, NHMRC Practitioner Fellowship.

Prof. Hu, Prof. Mackay and Dr. Griffanti wish to thank the Monument Trust Discovery Award from Parkinson’s UK, the MRC Dementias Platform UK and the National Institute for Health Research (NIHR) Oxford Biomedical Research Centre (BRC), and the NIHR Oxford Health BRC.

Prof. Seok Jong Chung wishes to thank the National Research Foundation of Korea funded by the Ministry of Education (Grant NRF-2018R1D1A1B07048959).

Dr. Rimona Weil wishes to thank the Wellcome Trust and the UCLH Biomedical Research Centre.

Prof. Henry Mak wishes to thank the State Key Laboratory of Brain and Cognitive Sciences, The University of Hong Kong.

## Supplemental Information

### S1. The ComBat harmonisation method and its application on this dataset

As noted in the Methods section, this algorithm was first developed for genomics analyses requiring to pool together data from different datasets (Johnson, Li and Rabinovic, 2007) and has been recently adapted to other measures, such as cortical thickness (Fortin et al., 2018). ComBat applies empirical Bayes (EB) estimation to account for site-specific variance by using an error with a multiplicative scaling factor that is scanner specific. The model allows to retain biological variance, such as that related to age, gender and disease (in our case, hallucinators and non hallucinators), variables that are entered the model as a vector. The algorithm is used on segmented and parcellated data, before running any other statistical analysis and has proven extremely reliable for this type of analyses as described in Fortin et al., 2018 and more recently by Radua et al., 2020. Hierarchical Bayes (HB) is also being proposed to harmonise data from different sites (Kia et al., 2020^1^). However, we chose to use EB and as the purpose of this study was not to assess the suitability of harmonisation models but to build on such harmonisation to develop models for the understanding of the mechanisms underlying visual hallucinations in PD. Secondly, EB is less dependent on the hyper-parameter settings and thus uses data more directly, which for our purposes was the most suitable choice, as HB would possibly be more suitable for approaches whereby a careful specification of hyper-parameters is crucial, as is the case for normative modelling (Kia et al., 2020^1^).

**Figure S1.**
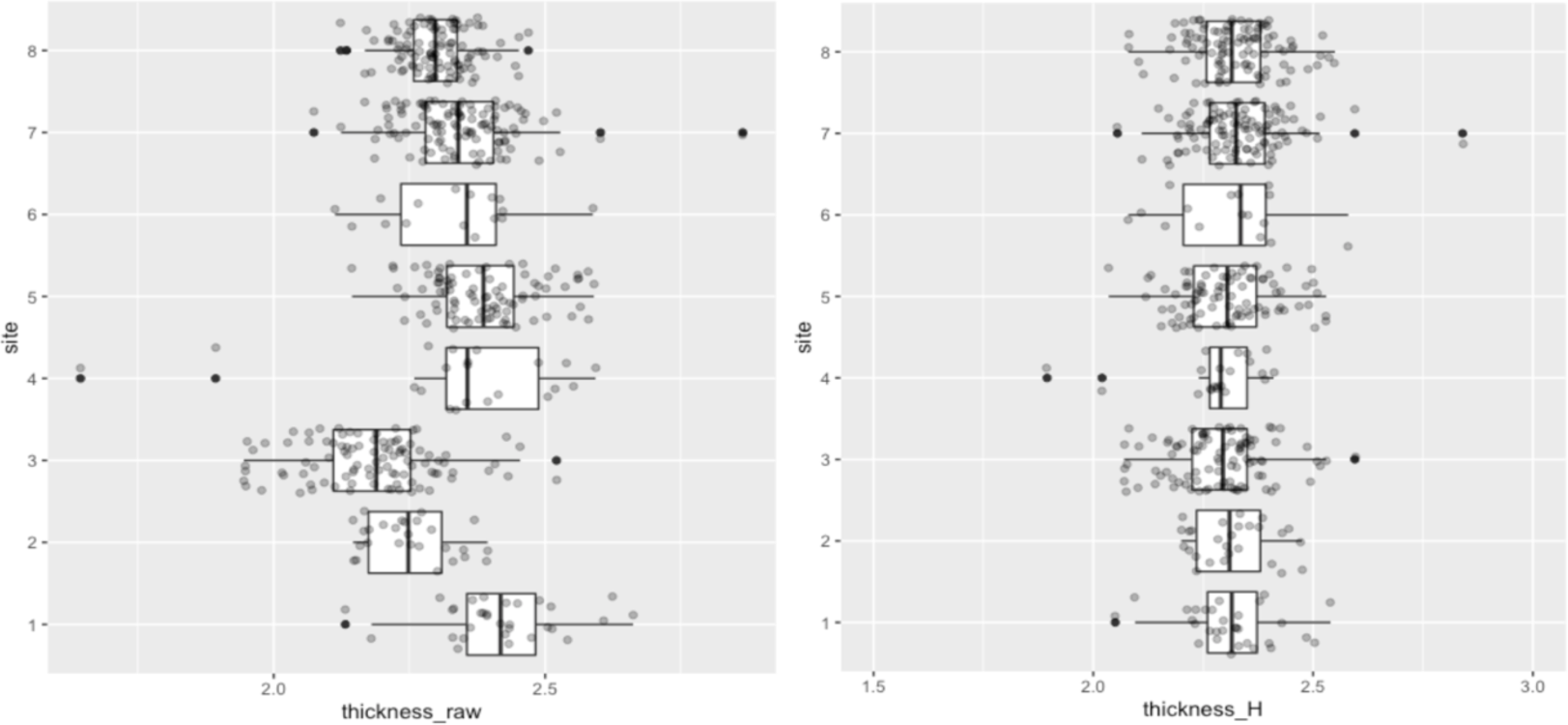
Mean thickness before (thickness_raw) and after harmonisation (thickness_H). On the X axis thickness values, on the Y axis the numbers correspond to the different sites. Sites: 1 = University of Lille; 2 = Hong Kong University; 3 = Yonsei University; 4 = Newcastle University; 5 = University of Sydney; 6 = KCL (Dr. ffytche); 7 = Oxford Discovery Cohort; 8 = University College London.

### S2. Additional information on the full sample

*Meta-analysis to determine whether participants in the full sample (N=493) are matched on the relevant demographic and clinical variables.* As we did not have raw data for all groups for all relevant variables, we tested whether our participants were overall matched for age, onset, MMSE, UPDRS and LED. The analysis was carried out with R package ‘metafor’. Results show that participants are overall matched for age, PD motor severity as assessed by the UPDRS-III total score, onset and levodopa equivalent daily dose (LED). The model shows a trend towards significant driven by the KCL dataset (N= 15) for the MMSE variable.

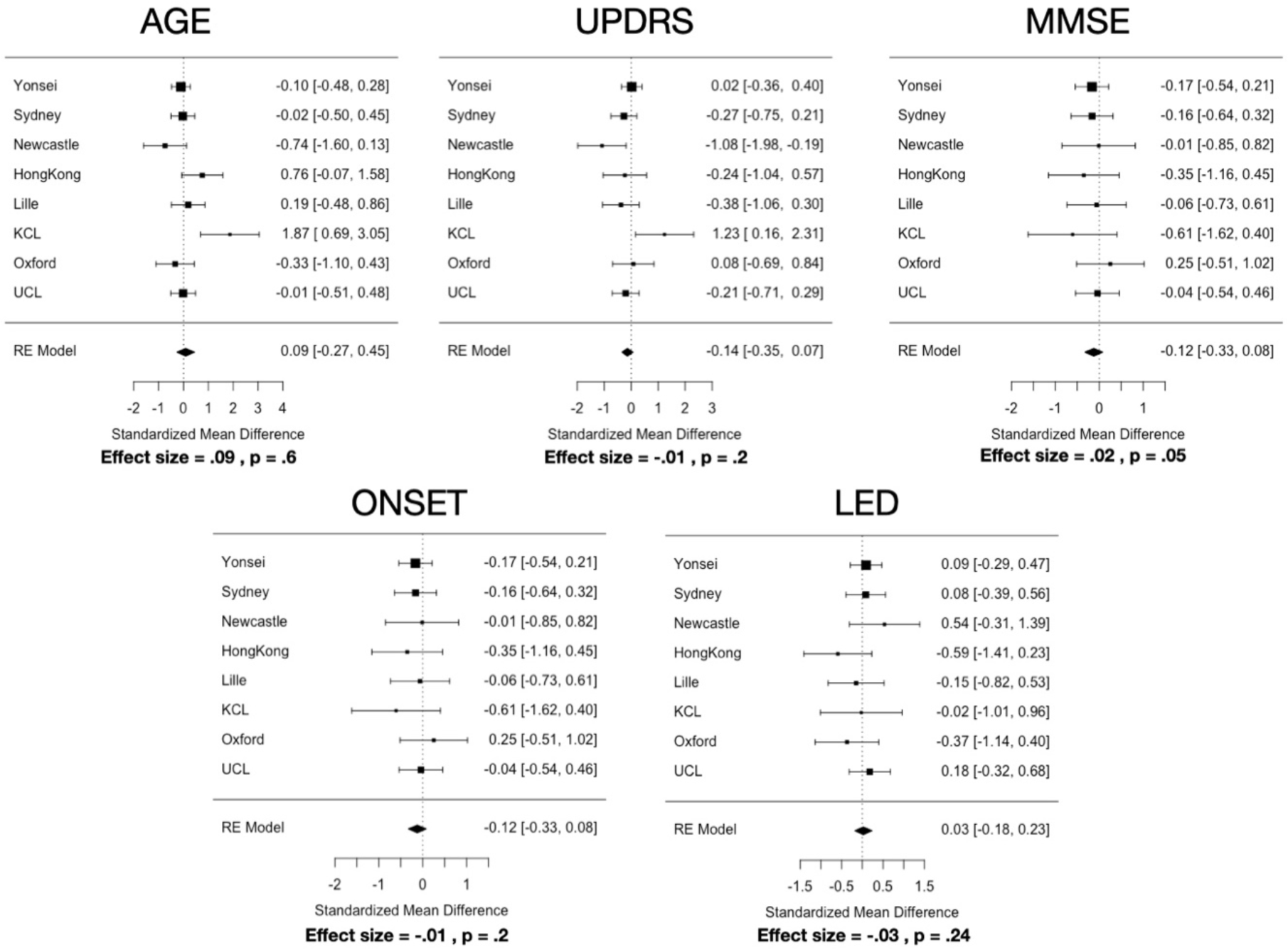

*Information on values for brain volume, total intracranial volume (TIV), total gray matter volume (GM) and age differences in the full sample.* Brain vol. (seg) describes the volume of all voxels in the aparc+aseg.mgz based on which the morphometric information were extracted. This includes voxels that are not background or brainstem, and includes vessel, optic chiasm and CSF segmentations. PD-VH did not significantly differ in TIV from PD-noVH (PD-VH, 1488363.39 ± 192038.64, PD-noVH, 1512273.57 ± 216293.33, p = .84), in the segmented brain volume (PD-VH =0.74 ± 0.05, PD-noVH = 0.74± 0.06, p = .12) but did differ in terms of gray matter volume (PD-VH 559525.16± 63591.98, PD-noVH 584869.68± 71580.08, p = .02).

### S3. Table for thickness, surface area and volume differences in PD with VH vs. PD no VH patients. Regions are reported in descendent order of significance (p value)

**Cortical thickness MANCOVA (covariates: age, gender, TIV)**
| *Region* | *df* | *Mean Square* | *F* | *p* | *Partial Eta Squared* |
| --- | --- | --- | --- | --- | --- |
| R_precuneus | 1, 488 | 0.456 | 16.330 | 0.000 | 0.032 |
| L occipitotemporal | 1, 488 | 0.402 | 15.570 | 0.000 | 0.031 |
| R_planumtemp | 1, 488 | 0.589 | 14.878 | 0.000 | 0.030 |
| L_sup_par | 1, 488 | 0.397 | 14.624 | 0.000 | 0.029 |
| R_ips | 1, 488 | 0.246 | 13.016 | 0.000 | 0.026 |
| R_parietalS | 1, 488 | 0.318 | 12.189 | 0.001 | 0.024 |
| L_ips | 1, 488 | 0.207 | 11.994 | 0.001 | 0.024 |
| R_occAnt_sul | 1, 488 | 0.394 | 11.724 | 0.001 | 0.023 |
| L_STempSul | 1, 488 | 0.195 | 11.464 | 0.001 | 0.023 |
| L_occAnt_sulc | 1, 488 | 0.424 | 11.420 | 0.001 | 0.023 |
| R_supTempsul | 1, 488 | 0.203 | 11.344 | 0.001 | 0.023 |
| L_precuneus | 1, 488 | 0.307 | 10.744 | 0.001 | 0.022 |
| L_ifg_orb | 1, 488 | 0.477 | 10.620 | 0.001 | 0.021 |
| R_occipitotemporal | 1, 488 | 0.291 | 10.614 | 0.001 | 0.021 |
| R_insula (short) | 1, 488 | 0.929 | 10.431 | 0.001 | 0.021 |
| R_ifgTr | 1, 488 | 0.321 | 10.421 | 0.001 | 0.021 |
| R_frontopolTrs | 1, 488 | 0.309 | 10.217 | 0.001 | 0.021 |
| R_mfg | 1, 488 | 0.194 | 10.107 | 0.002 | 0.020 |
| L_sfg | 1, 488 | 0.229 | 9.913 | 0.002 | 0.020 |
| L_itg | 1, 488 | 0.338 | 9.699 | 0.002 | 0.019 |
| L_cingMarg_s | 1, 488 | 0.240 | 9.647 | 0.002 | 0.019 |
| L_ifg_op | 1, 488 | 0.234 | 9.454 | 0.002 | 0.019 |
| L_inf_angular | 1, 488 | 0.244 | 9.385 | 0.002 | 0.019 |
| R_precentrInf | 1, 488 | 0.230 | 9.248 | 0.002 | 0.019 |
| L_temp_pole | 1, 488 | 0.506 | 9.193 | 0.003 | 0.018 |
| R_subcall | 1, 488 | 1.222 | 8.688 | 0.003 | 0.017 |
| L_cuneus | 1, 488 | 0.159 | 8.579 | 0.004 | 0.017 |
| R_cuneus | 1, 488 | 0.164 | 8.030 | 0.005 | 0.016 |
| L_parietoOc | 1, 488 | 0.156 | 7.819 | 0.005 | 0.016 |
| R_occsupG | 1, 488 | 0.209 | 7.230 | 0.007 | 0.015 |
| R_octemp | 1, 488 | 0.123 | 7.086 | 0.008 | 0.014 |
| R_supFr | 1, 488 | 0.134 | 7.000 | 0.008 | 0.014 |
| R_latFissureP | 1, 488 | 0.152 | 6.977 | 0.009 | 0.014 |
| R_sfg | 1, 488 | 0.144 | 6.728 | 0.010 | 0.014 |
| R_ifg_op | 1, 488 | 0.202 | 6.680 | 0.010 | 0.014 |
| R_mtg | 1, 488 | 0.218 | 6.541 | 0.011 | 0.013 |
| R_infang | 1, 488 | 0.153 | 6.391 | 0.012 | 0.013 |
| R_subpar | 1, 488 | 0.203 | 6.344 | 0.012 | 0.013 |
| L_stg_lat | 1, 488 | 0.276 | 6.341 | 0.012 | 0.013 |
| L_supramarg. Inf | 1, 488 | 0.164 | 6.317 | 0.012 | 0.013 |
| R_itg | 1, 488 | 0.207 | 6.241 | 0.013 | 0.013 |
| L_occ_s | 1, 488 | 0.198 | 6.197 | 0.013 | 0.013 |
| L_plan_tempor_s | 1, 488 | 0.249 | 6.173 | 0.013 | 0.012 |
| L_rectus | 1, 488 | 0.203 | 6.007 | 0.015 | 0.012 |
| R_temporalPOle | 1, 488 | 0.357 | 5.977 | 0.015 | 0.012 |
| L_stg | 1, 488 | 0.299 | 5.682 | 0.018 | 0.012 |
| R_cing_marg | 1, 488 | 0.129 | 5.622 | 0.018 | 0.011 |
| R_latfusif | 1, 488 | 0.235 | 5.606 | 0.018 | 0.011 |
| L_ifg_triang | 1, 488 | 0.163 | 5.578 | 0.019 | 0.011 |
| L_ifs | 1, 488 | 0.098 | 5.560 | 0.019 | 0.011 |
| R_orbG | 1, 488 | 0.149 | 5.499 | 0.019 | 0.011 |
| R_precentr | 1, 488 | 0.316 | 5.472 | 0.020 | 0.011 |
| R_ifg_orb | 1, 488 | 0.301 | 5.354 | 0.021 | 0.011 |
| R_infTempsul | 1, 488 | 0.165 | 5.253 | 0.022 | 0.011 |
| R_precentS | 1, 488 | 0.159 | 5.248 | 0.022 | 0.011 |
| L_precentralg | 1, 488 | 0.285 | 5.203 | 0.023 | 0.011 |
| L_mfg | 1, 488 | 0.099 | 5.038 | 0.025 | 0.010 |
| R_moccG | 1, 488 | 0.125 | 5.016 | 0.026 | 0.010 |
| L_paracentral_g | 1, 488 | 0.197 | 4.893 | 0.027 | 0.010 |
| R_frontomarg | 1, 488 | 0.200 | 4.886 | 0.028 | 0.010 |
| L_tempTrsSul | 1, 488 | 0.301 | 4.877 | 0.028 | 0.010 |
| R_rectus | 1, 488 | 0.206 | 4.810 | 0.029 | 0.010 |
| L_fiss_hor_ant | 1, 488 | 0.270 | 4.777 | 0.029 | 0.010 |
| L_fis_post | 1, 488 | 0.112 | 4.758 | 0.030 | 0.010 |
| L_occ_m | 1, 488 | 0.116 | 4.718 | 0.030 | 0.010 |
| R_calcarine | 1, 488 | 0.084 | 4.558 | 0.033 | 0.009 |
| R_supTrans | 1, 488 | 0.103 | 4.441 | 0.036 | 0.009 |
| L_sfs | 1, 488 | 0.090 | 4.356 | 0.037 | 0.009 |
| R_stglat | 1, 488 | 0.187 | 4.276 | 0.039 | 0.009 |
| L_octempSul_lat | 1, 488 | 0.158 | 4.263 | 0.039 | 0.009 |
| R_octemp_Lat | 1, 488 | 0.162 | 4.205 | 0.041 | 0.009 |
| L_trsv | 1, 488 | 0.096 | 4.195 | 0.041 | 0.009 |
| L_collaTr_ants | 1, 488 | 0.186 | 4.184 | 0.041 | 0.009 |
| L_central | 1, 488 | 0.070 | 4.100 | 0.043 | 0.008 |
| L_ins_g_short | 1, 488 | 0.298 | 4.068 | 0.044 | 0.008 |
| R_orbH | 1, 488 | 0.151 | 4.067 | 0.044 | 0.008 |
| R_mfs | 1, 488 | 0.079 | 4.063 | 0.044 | 0.008 |
| R_inffrontsulc | 1, 488 | 0.069 | 3.883 | 0.049 | 0.008 |
| R_paracentr | 1, 488 | 0.152 | 3.872 | 0.050 | 0.008 |
| R_latfissAntvert | 1, 488 | 0.320 | 3.836 | 0.051 | 0.008 |
| R_parietoOcc | 1, 488 | 0.067 | 3.724 | 0.054 | 0.008 |
| R_supramarg inf. | 1, 488 | 0.097 | 3.514 | 0.061 | 0.007 |

\* in red trends are reported

Surface area *MANCOVA with gender, age and TIV as covariates*
| *Region* | *df* | *Mean squared* | *F* | *p* | *Partial Eta Squared* |
| --- | --- | --- | --- | --- | --- |
| R_occipitotemporal | 1, 488 | 796730.837 | 10.876 | 0.001 | 0.022 |
| L_frontomarginal | 1, 488 | 93786.402 | 8.700 | 0.003 | 0.018 |
| L_insula Ig+central | 1, 488 | 29582.402 | 6.324 | 0.012 | 0.013 |
| L_precentral | 1, 488 | 119647.002 | 3.888 | 0.049 | 0.008 |
| R_orbitl lat sulc | 1, 488 | 16649.104 | 3.835 | 0.051 | 0.008 |
| L_subcallosal | 1, 488 | 61304.202 | 3.776 | 0.053 | 0.008 |
| L_lat fiss (ant) | 1, 488 | 6391.183 | 3.682 | 0.056 | 0.007 |
| L_insula circ. | 1, 488 | 12820.027 | 3.602 | 0.058 | 0.007 |
| L_calcarine | 1, 488 | 357969.184 | 3.489 | 0.062 | 0.007 |

\* in red trends are reported

| *Region* | *df* | *Mean squared* | *F* | *p* | *Partial Eta Squared* |
| --- | --- | --- | --- | --- | --- |
| lh_G_front_inf-Orbital_area | 1, 489 | 13656.353 | 5.125 | 0.024 | 0.010 |
| rh_G_front_middle_area | 1, 489 | 851980.733 | 5.423 | 0.020 | 0.011 |
| Rh_occipital_pole | 1, 489 | 355496.685 | 4.652 | 0.032 | 0.009 |
| rh_G_front_sup_area | 1, 489 | 1155724.296 | 4.431 | 0.036 | 0.009 |
| rh_calcarine_S | 1,489 | 448355.197 | 5.007 | 0.026 | 0.010 |
| lh_calcarine_S | 1,489 | 541601.042 | 5.165 | 0.023 | 0.010 |
| rh_G&S_occipital_inf_area | 1, 489 | 70740.397 | 3.652 | 0.057 | 0.007 |

*Subcortical volumes MANCOVA with age, gender and TIV as covariates*
| *Region* | *df* | *Mean squared* | *F* | *p* | *Partial Eta Squared* |
| --- | --- | --- | --- | --- | --- |
| Left-Cerebellum-WM | 1, 488 | 25136509.462 | 4.803 | 0.029 | 0.010 |
| L amygdala | 1, 488 | 349741.740 | 6.664 | 0.010 | 0.013 |
| R amygdala | 1, 488 | 278036.600 | 5.335 | 0.021 | 0.011 |

### S4. NPI subgroup custom MANCOVAs with binary score (PD-VH vs. PD-noVH) and correlation analysis (VH only) with LED as covariate

The NPI subgroup (146 patients, 79 PD-noVH, 67 PD-VH) is the group for which we had the richest clinical dataset, as for most of these patients we have not only age and gender, but also disease onset, levo-dopa equivalent daily dose (LED) and mini mental state examination (MMSE) data, PD severity, together with the continuous NPI score. 126 had UPDRS-III scores. 20 from the same group did not have those scores. We computed the mean for the group regardless of VH and used that score in *our* model.

Patients are matched for gender (35 females PD-noVH, 27 females PD-VH) (***χ****2* = .238, *p* = 0.62), age (PD-VH=70.39 ± 6.82, PD-noVH = 69.64 ± 6.45) [*F*(1,144) = 0.47, *MSE* = 20.39, *p* = .5], disease onset (PD-VH=5.86 ± 5.40, PD-noVH = 4.64 ± 5.22) [*F*(1,144) = 1.91, *MSE* = 53.85, *p* = .17], levodopa equivalent daily dose (LED) (PD-VH=575.86 ± 366.43 mg, PD-noVH =502.17± *327*.34 mg) [*F*(1,144) =1.60, *MSE* =191615.74, *p* = .21], and MMSE score (PD-VH=26.28±2.69, PD-noVH =26.81±2.51), [*F*(1,113)=1.49, *MSE* = 10.05, *p* = .22)].

Patients were matched for UPDRS-III within each study. However, when carrying out a one-way ANOVA using the mean to fill the 20 missing values (N PD-VH =7, N PD-noVH = 13) they were not matched for UPDRS-III, with PD-VH having a higher score [*F*(1,113)=6.9, *MSE* = 1187.07, *p* = .01)]. We ran a custom multivariate ANOVA model with MMSE, gender, age, TIV, UPDRS-III, LED and onset as covariates, controlling *also* for the interaction of those covariates that correlated with each other (age * LED + age * MMSE + onset * LED + onset * UPDRS3 +LED * MMSE).

**Table S4.** Demographics. First, we present a summary of the demographics and clinical information of this subsample; for UPDRS-III values, we report the scores including the N=20 subjects with missing values; for original UPDRS-III scores for each group see **Table 1** in the main text (study: Yonsei University; University of Newcastle; University of Lille). **Cortical thickness.** Multivariate analysis results for cortical thickness differences in PD+VH vs. PD noVH patients. **Surface area.** Multivariate analysis results for SA differences in PD+VH vs. PD noVH patients. **Subcortical volumes.** Multivariate analysis results for subcortical volume differences in PD+VH vs. PD noVH patients. Regions are sorted by p value. * values greater for PD with VH; * trends are indicated in red.

**Demographics**
| *Group* | *Age* | *MMSE* | *LED* | *Onset* | *UPDRS-III* |
| --- | --- | --- | --- | --- | --- |
| PD-VH<br>(N=67, 27 F) | 70.39<br>(SD=6.82) | 26.28 (SD<br>=2.69) | 575.86 mg (SD<br>=366.43) | 5.86 (SD<br>=5.40) | 30.32 (SD<br>=15.25) |
|  | *p* = .49 | *p* = .22 | *p* = .21 | *p* = .17 | *p* = .01 |
| PD-noVH<br>(N=79, 35 F) | 69.64 (SD<br>=6.45) | 26.81(SD<br>=2.51) | 503.17 mg (SD<br>=327.34) | 4.64 (SD<br>=5.22) | 24.61 (SD<br>=10.93) |

**Cortical thickness**
| *Region* | *df* | *Mean squared* | *F* | *p* | *Partial Eta Squared* |
| --- | --- | --- | --- | --- | --- |
| r planum tempo | 1,145 | 0.59 | 16.96 | 0.00 | 0.98 |
| L cing marg | 1,145 | 0.29 | 13.75 | 0.00 | 0.96 |
| R STS | 1,145 | 0.22 | 13.01 | 0.00 | 0.95 |
| R subcentral | 1,145 | 0.38 | 12.74 | 0.00 | 0.94 |
| r SMG | 1,145 | 0.28 | 10.61 | 0.00 | 0.90 |
| R IPS | 1,145 | 0.18 | 10.37 | 0.00 | 0.89 |
| L STS | 1,145 | 0.17 | 10.35 | 0.00 | 0.89 |
| L occtemp ML | 1,145 | 0.24 | 10.30 | 0.00 | 0.89 |
| L sup par | 1,145 | 0.26 | 10.02 | 0.00 | 0.88 |
| R jensen | 1,145 | 0.38 | 9.41 | 0.00 | 0.86 |
| L tempo pole | 1,145 | 0.45 | 9.29 | 0.00 | 0.86 |
| L coll tran ant | 1,145 | 0.37 | 9.04 | 0.00 | 0.85 |
| r occtemp ML | 1,145 | 0.21 | 8.70 | 0.00 | 0.83 |
| R ITS | 1,145 | 0.25 | 8.20 | 0.00 | 0.81 |
| r parietal sucl | 1,145 | 0.20 | 8.04 | 0.01 | 0.80 |
| L IPS | 1,145 | 0.13 | 7.75 | 0.01 | 0.79 |
| r precuneus | 1,145 | 0.18 | 7.66 | 0.01 | 0.78 |
| L IFS | 1,145 | 0.12 | 7.62 | 0.01 | 0.78 |
| R MTG | 1,145 | 0.23 | 7.40 | 0.01 | 0.77 |
| R precentral inf | 1,145 | 0.18 | 7.28 | 0.01 | 0.76 |
| R m occ g | 1,145 | 0.17 | 7.10 | 0.01 | 0.75 |
| L oocctemp sulc | 1,145 | 0.27 | 7.09 | 0.01 | 0.75 |
| R occ ant sulc | 1,145 | 0.21 | 6.68 | 0.01 | 0.73 |
| R lat fiss | 1,145 | 0.14 | 6.58 | 0.01 | 0.72 |
| R trans post | 1,145 | 0.26 | 6.57 | 0.01 | 0.72 |
| R cuneus | 1,145 | 0.12 | 6.28 | 0.01 | 0.70 |
| L fiss hor ant | 1,145 | 0.32 | 6.18 | 0.01 | 0.69 |
| R IFS | 1,145 | 0.11 | 6.15 | 0.01 | 0.69 |
| R ifg op | 1,145 | 0.19 | 6.05 | 0.02 | 0.68 |
| R ifg orb | 1,145 | 0.38 | 5.95 | 0.02 | 0.68 |
| L _paracentral_g | 1,145 | 0.24 | 5.91 | 0.02 | 0.67 |
| L precuneus | 1,145 | 0.16 | 5.89 | 0.02 | 0.67 |
| r inf angular | 1,145 | 0.12 | 5.80 | 0.02 | 0.67 |
| L occ ant sulc | 1,145 | 0.20 | 5.63 | 0.02 | 0.65 |
| r ITG | 1,145 | 0.17 | 5.61 | 0.02 | 0.65 |
| R ifg tri | 1,145 | 0.18 | 5.59 | 0.02 | 0.65 |
| R parietocc | 1,145 | 0.09 | 5.53 | 0.02 | 0.65 |
| L ITG | 1,145 | 0.18 | 5.46 | 0.02 | 0.64 |
| L planum polar<br>sup | 1,145 | 0.45 | 5.30 | 0.02 | 0.63 |
| L coll tr post | 1,145 | 0.18 | 5.14 | 0.02 | 0.61 |
| R occtemplat fus | 1,145 | 0.20 | 5.13 | 0.03 | 0.61 |
| L ITS | 1,145 | 0.13 | 4.97 | 0.03 | 0.60 |
| R MFG | 1,145 | 0.10 | 4.94 | 0.03 | 0.60 |
| r insula sup | 1,145 | 0.11 | 4.87 | 0.03 | 0.59 |
| L precentral | 1,145 | 0.26 | 4.83 | 0.03 | 0.59 |
| L occ m | 1,145 | 0.11 | 4.72 | 0.03 | 0.58 |
| L lunauts | 1,145 | 0.11 | 4.63 | 0.03 | 0.57 |
| R subpar | 1,145 | 0.12 | 4.53 | 0.04 | 0.56 |
| L ifg triang | 1,145 | 0.12 | 4.49 | 0.04 | 0.56 |
| L jensen | 1,145 | 0.44 | 4.42 | 0.04 | 0.55 |
| L ifg orb | 1,145 | 0.18 | 4.32 | 0.04 | 0.54 |
| R frontpolar | 1,145 | 0.12 | 4.30 | 0.04 | 0.54 |
| L suborb | 1,145 | 0.27 | 4.21 | 0.04 | 0.53 |
| L parietocc | 1,145 | 0.09 | 4.09 | 0.05 | 0.52 |
| L SMG | 1,145 | 0.10 | 4.00 | 0.05 | 0.51 |
| R temporal pole | 1,145 | 0.21 | 3.93 | 0.05 | 0.50 |
| R oocc inf | 1,145 | 0.19 | 3.92 | 0.05 | 0.50 |
| L_ifg_op | 1,145 | 0.08 | 3.58 | 0.06 | 0.47 |
| R Sup transv | 1,145 | 0.09 | 3.57 | 0.06 | 0.47 |
| L insula ant | 1,145 | 0.17 | 3.55 | 0.06 | 0.46 |
| L sfg | 1,145 | 0.07 | 3.50 | 0.06 | 0.46 |
| r precentral | 1,145 | 0.20 | 3.50 | 0.06 | 0.46 |

**Surface Area**
| **Region** | **df** | **Mean squared** | **F** | **p** | **Partial Eta Squared** |
| --- | --- | --- | --- | --- | --- |
| lh_S_precentral-inf-part_area | 1,145 | 149160.25 | 6.22 | 0.01 | 0.70 |
| lh_G_Ins_lg&S_cent_ins | 1,145 | 18462.45 | 5.49 | 0.02 | 0.64 |
| lh_S_circular_insula_sup | 1,145 | 55107.36 | 5.11 | 0.03 | 0.61 |
| rh_G_oc-temp_med-Lingual_area | 1,145 | 266848.02 | 4.59 | 0.03 | 0.57 |
| rh_S_oc_middle&Lunatus_area | 1,145 | 56153.57 | 3.39 | 0.07 | 0.45 |

| **Region** | **df** | **Mean squared** | **F** | **p** | **Partial Eta Squared** |
| --- | --- | --- | --- | --- | --- |
| L amygdala | 1,145 | 240574.72 | 7.77 | 0.01 | 0.79 |
| L Hippocampus | 1,145 | 721840.97 | 6.68 | 0.01 | 0.73 |
| WM hypointensities | 1,145 | 59267855.43 | 5.46 | 0.02 | 0.64 |
| R caudate | 1,145 | 844168.41 | 4.67 | 0.03 | 0.57 |
| R hippocampus | 1,145 | 582591.89 | 3.73 | 0.06 | 0.48 |

For thickness, no significant main effect of gender, age, TIV, MMSE, LED, UPDRS, onset, age * LED + age * MMSE + onset * LED + onset * UPDRS-III + LED * MMSE was observed.

For SA, no significant main effect of gender, age, TIV, MMSE, LED, UPDRS, onset, age * LED + age * MMSE + onset * LED + onset * UPDRS-III + LED * MMSE was observed.

For subcortical volumes, we observed a main effect of age* LED [F(1,115)=1.53, p =.04], onset* LED [F(1,115)=1.66, p=.02], onset* UPDRS-III [F(1,115)=1.70, p=.02], gender [F(1,115)=3.23, p<.001], onset [F(1,115)=1.73, p=.01], TIV [F(1,115)=2584.07, p<.001], UPDRS-III [F(1,115)=1.82, p =.008], but not of MMSE, LED, age, LED*MMSE, age * MMSE.

Finally, we carried out the same correlational analysis described in the main text, but with LEDD as a covariate, as it might be related to severity of VH, obtaining the same results: negative correlations were found for the right superior temporal sulcus (*r* = -.26, *p* = .03), the right inferior parietal sulcus (*r* = −.24, *p* = .05), the right Jensen sulcus (*r* = -.28, *p* = .02) and the right cingulum marginalis (*r* = -.24, *p* = .056), which was a trend in this case. In addition there was the positive correlation reported also in the main text with the right frontomarginal gyrus (*r* = .26, *p* = .04).

### S5. Subgroup analysis with ordinal VH score

An additional sensitivity analysis was carried out for a large subgroup, that is all those patients for which we had additional information about the degree of hallucinations, levodopa equivalent dose (LED) and onset date. Overall, the 128 PD-VH patients (64 mild, 47 moderate, 17 severe) were matched for age [F(2,125)=1.59, p=0.21], onset [F(2,125)=0.18, p=0.84], gender (*X^2^*=.87, p=.65) and TIV [F(2,125)=0.07, p=0.93] (we did not have enough LED or cognition data to check for these variables).

Patients with mild VH had a mean age of 66.71*±* 8.20, those with moderate VH 69.10 *±* 7.37 and those with severe VH 69.34 *±* 7.30. Those with mild VH had a disease duration of 5.29*±* 4.06 years, those with moderate VH 5.57*±* 5.58 and those with severe 4.79 *±* 4.05. Finally, patients with mild VH had a mean TIV of 1477284.73 *±*178401.32, patients with moderate VH 1490485.07 *±*188476.12, patients with severe VH 1480473.38 *±*195998.02.

As there is not a specific criterion to do so, in order to create an ordinal variable, for patients we had UPDRS scoring we have retained that (0 = no hallucinations or delusions, 1= illusions or non formed hallucinations, 2 = Formed hallucinations independent of environmental stimuli, 3= Formed hallucinations with loss of insight, 4= patient has delusions or paranoia (but no patients had a UPDRS 4 score or a NPI, NEVHI, MIAMI equivalent). We have “translated” the NPI continuous score to such a scale, with scores ranging from 1 to 3 categorised as “minor/mild VH”, scores raging from 4 to 8 as “moderate” and scores above 9 as “severe”; for the MIAMI we *have* used a similar procedure, as the scale ranges from 0 to 14. Finally, for the NEVHI, we have used the raw data from the interviews to determine whether the person had mild, moderate or severe visual hallucinations.

To isolate regions where there was some difference between the groups in order to create a multi-region ordinal regression model, we ran a multivariate ANOVA for each morphometric measure (subcortical volumes, SA, cortical thickness) was ran with *degree of hallucinations as between subjects factor* on hallucinators only (McCrum-Gardner, 2008) and with age, onset, gender, TIV as covariates.

For thickness we used age [F(2,121)=1.25, p=0.62], gender[F(2,121)=1.95, p=0.52], onset [F(2,121)=.3.91, p=0.37], and TIV[F(2,121)=.455, p=.86], as covariates, finding no main effect for any of these factors except for TIV. Participants differed in thickness in the left temporal pole, left insula, left collateralis transversalis posterior, left suborbital gyrus, right frontomarginal gyrus (as in the NPI correlational analysis) and in the right lateral horizontal fissure, with severe and moderate hallucinators having higher thickness in these regions. We speculate *that* greater thickness in these regions may constitute a pre-existing risk factor for aggravation of VH severity but additional data would be needed to further explore this hypothesis.

For SA the covariates age [F(2,121)=.42, p=0.87], gender[F(2,121)=5.25, p=0.34], onset [F(2,121)=.88, p=0.68], TIV [F(2,121)=4.64, p=0.36] showed no main effect. Participants differed in SA in the occipitotemporal gyri and in the precuneus bilaterally, calcarine fissure, insula and precentral gyrus; SA was reduced in severe hallucinators if compared to mild and moderate hallucinators.

For subcortical volumes we found a main effect of age [F(2,121)=2.91, p<.001], gender[F(2,121)=2.02, p=0.003], and TIV[F(2,121)=769.44, p<.001], but not of onset [F(2,121)=1.37, p=0.11]. Participants differed in volume in the bilateral inferior lateral ventricles, with moderate hallucinators having greater ventricle volume than mild hallucinators.

We entered the regions emerged as significant in ordinal logistic regression models carried out with R package MASS. For *thickness*, the intercepts for mild vs. moderate hallucinators (*t* =1.87, *st.error* = 3.41, *intercept* = 6.37, *p*=.06) showed a trend towards significant and was significant for moderate vs. severe (*t* =2.42, *st.error* = 3.45, *intercept* = 8.36, *p* =.01), were significant with *residual deviance* = 241.43, *AIC* = 257.43. Interestingly, thickness in these regions is greater in the moderate and severe hallucinators overall, recalling how the right frontomarginal thickness positively correlated with the NPI score in the smaller sample. However, none of the regions significantly predicted the hallucinations ordinal score (all *t* values < ±1.5 and all *ps*>.05).

For *surface area* mild vs. moderate hallucinators (*t* = -1529.65, *st.error* = 0.001, *intercept* = -1.71, *p* <.0001) was significant, but not for moderate vs. severe (*t* =1.44, *st.error* = 0.28, *intercept* = 0.41, *p* =.15), were significant with *residual deviance* = 231.47, *AIC* = 253.47, with the coefficients for the left occipitotemporal fusiform gyrus being above the ∼2 threshold for the t value (*t* =-2.33, *p* =.001) and being greater in mild vs. moderate, and the left insula circularis showing the opposite pattern (*t* =2.87, *p* =.004). The lack of significance when considering the comparison with those with a ‘severe’ score may be due to different reasons, with one being that the different PDP scales used in the different studies have slightly different criteria, but also that patients with higher scores may have delusions as well, which may as well be in the same continuum as visual hallucinations, but this does not necessarily imply a graver atrophy in the same regions underlying VH. Finally, the three groups have very different sample sizes, and this *might* as well affect the analyses, as one can notice the large variability within each group.

### S6. Additional models and details about receptor density maps

#### a) Additional models for thickness and surface area

We used bootstrapping to estimate the confidence intervals of the *regression* models that resulted significant and that are reported in the main text.

For *cortical thickness* the estimation of the CI of the *significant* regression model with 5-HT_2A_ as predictor and the difference of the means between PD-VH and PD-noVH for regions which differed between groups (*β* =-.252, *t* = -2.2, *p* =.03) shows the model for the coefficients was significant *p* = .004, CI (-.0003,-.00005). A similar pattern is observed for 5-HT_1A_ (*β* = -.26, *t* =-2.25, *p* = 0.03), with bootstrapping returning a significant model for the coefficients was significant *p* = .005, CI (-.0003, -.00006) and for the D2/D3 model *(β* =-.35, *t* =-3.14, *p* = 0.002), with *p* <.001, CI (-.67, -.38).

For *surface area*, as mentioned in the Methods, we used regions that resulted different in PD-VH vs. PD-noVH from an exploratory one-way ANOVA, as the number of regions resulting different in the basis of the MANCOVAs performed and reported in S3 were too small in number to carry out a well powered model. We estimated the CI of the regression model with 5-HT_2A_ as predictor and the difference of the means between PD-VH and PD-noVH for regions which differed between groups (*β* =-.22, *t* = 2.1, *p* =.038), finding that the model for the coefficients was significant *p* = .003, CI (.08, .41). For the same *significant* regression model with 5-HT_1A_ (*β* =.27, slope= 0.22, *t* = 2.2, *p* =.01), we found that the model for the coefficients was significant *p* = .012, CI (.09, .35). For the D2/D3 model (*β* = .31, *t* = 3.038, *p* = .003) the estimation of the CI was significant, *p* < .001, CI (112.8, 617.7).

In addition, since both 5-HT_1A_ (*β* =.181, *t* = 2.07, *p* = .04) and D2/D3 (*β* =.277, *t* = 3.24, *p* = .001) resulted as significant predictors also when considering all regions, we repeated the same procedure, finding that the bootstrap for the coefficients resulted significant for 5-HT_1A_, *p* = .008, CI (0.06, .40) and for D2/D3 *p* < .001, CI (180.08, 516.77).

For regions where there was a relationship between receptor density and structural morphometrics for all regions, additional analyses were conducted to assess whether this was simply the same effect as the relationship of the regions that differed or a feature of the whole brain by examining the non-significant regions.

*Cortical Thickness.* The models with 5-HT_2A_ binding potential per region as predictor and the mean difference per region (considering only the regions where no difference between groups was observed) as dependent variable resulted not *significant* for regions that did not differ (*β* = -.013, t=-.09 p=0.93); a similar result was obtained for 5-HT_1A_ (*β* =.05, t=.35, p= 0.72) and D2/D3 (*β* =.24, t=1.72, p= .09) for the same regions.

*Surface Area*. The models with 5-HT_2A_ binding potential per region as predictor and the mean difference per region (considering only the regions where no difference between groups was observed) as dependent variable resulted not significant for regions that did not differ (*β* =-.013, *t* = -.067 *p* = .5); a similar result was found for 5-HT_1A_ (*β* =-.05, *t* = -1.18, *p* = .3), whereas D2/D3 was a significant predictor also for this model (*β* =.243, *t* =2.91, *p* =.001).

*Slope comparisons*. For thickness we compared the slopes for 5-HT_2A_, 5-HT_1A_ and D2/D3 for the differing regions, finding that the three slopes did not differ (*p*> .05). For surface area we observed a similar result (p > .05).

#### b) Receptors binding potentials models for volumes models results

Regression models were carried out to estimate the relationship between subcortical volumes (N=19) mean differences between hallucinators and non-hallucinators and receptor density profiles. As significantly differing regions were just the hippocampi and the amygdala, we ran separate models for each receptor using all regions’ subcortical volumes. The models resulted not significant when 5-HT_2A_ binding potential per region was used as predictor (*β = -*.14*, t* = -0.6, *p* = .5), when 5-HT_1A_ (*β = -*.24*, t* = -1.01, *p* = .32) and D2/D3 (*β* = .06*, t* = .24, *p* = .82) were used as predictors.

**c)** In order to better understand these results, we ran Pearson’s correlations between each receptor’s density maps at the cortical level, finding that 5-HT_2A_ and D2/D3 did now show a significant correlation (*r* =.15 *p* = .07), whereas 5-HT_2A_ and 5-HT_1A_ did (*r* =.92, p <.001) and also 5-HT_1A_ and D2/D3 (*r* =.27, *p* =.002).

### S7. PCA scree plots

**Figure 7.2.**
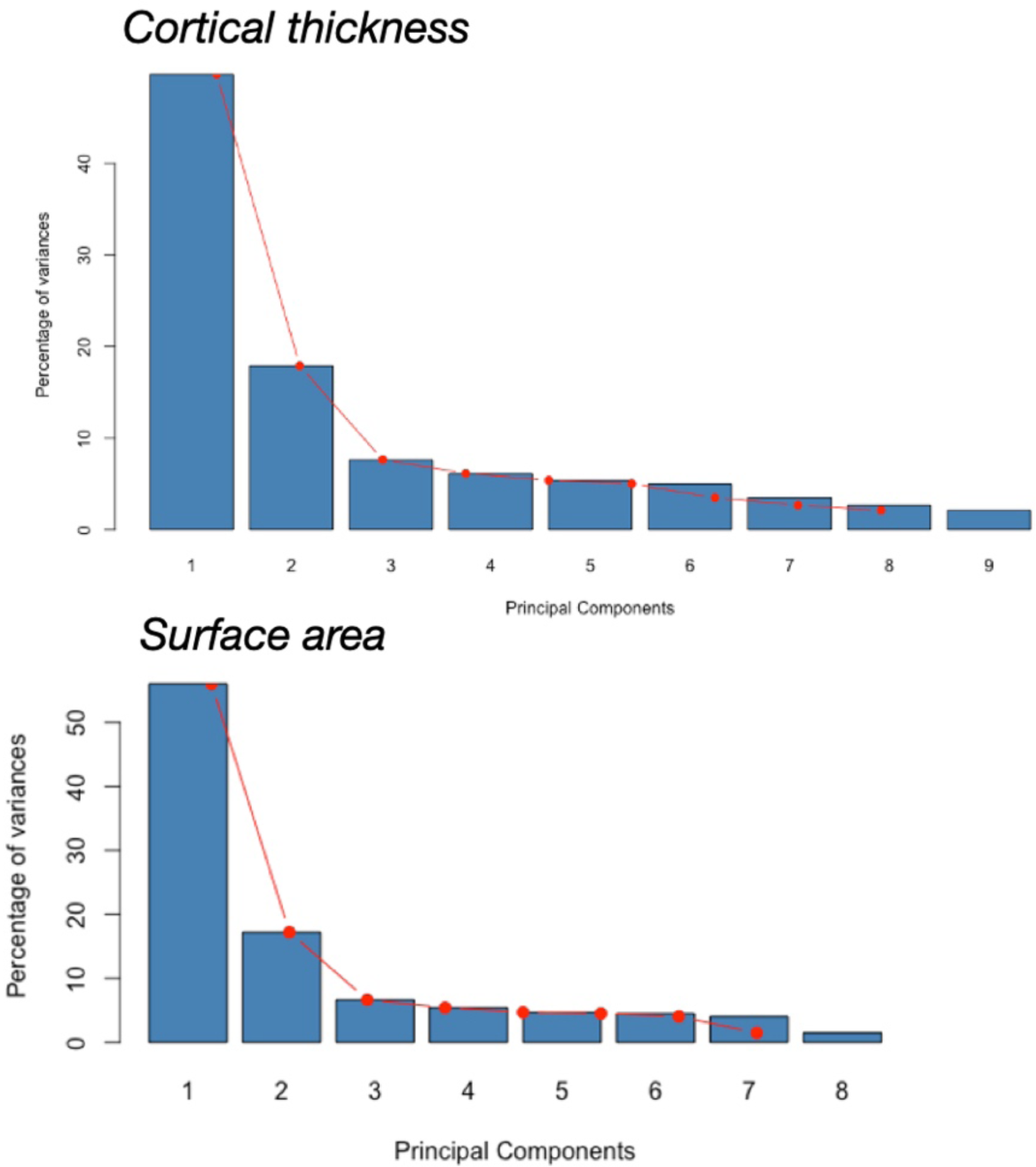
**a)** Scree Plot for Cortical thickness data. On the y axis, the % of variance explained by each component, on the y axis the components number. Following the criterion of picking only components with eigenvalues >1, PC 1 and 2 were selected.**b)** Scree Plot for Surface area data. On the y axis, the % of variance explained by each component, on the y axis the components number. Following the criterion of picking only components with eigenvalues >1, PC 1 and 2 were selected.

### S8. Structural covariance

Differences in inter-regional correlations between PD-VH and PD-noVH represented with a correlogram. Covariance matrices computed on the GLM residuals used for the structural covariance analysis were used to compute the difference in the inter-regional correlation coefficients in [PD-VH – PDnoVH] patients (**Figure S8**; those depicted are the difference in the coefficients). A cell by cell comparison of the correlation coefficients with the *cocor* package for R was computed to investigate regions that showed the highest covariance in PD-VH and in PD-noVH. In purple, inter-regional correlation coefficients that were greater for PD-VH, in yellow those greater for PD-noVH. The analysis on the z scores (Fischer) highlighted that the most significant regions with the most inter-regional correlation with other regions for VH patients were the left caudal MFG, left cuneus, left IPL, left ITG, left iCC, left LOG, left middle orbitofrontal gyrus, left paracentral gyrus, left IFG opercularis, left temporoparietal, right parahippocampal gyrus, right iFG opercularis, right IFG triangular, right temporoparietal. In particular the left cMFG highly correlated with parieto-occipital regions (IPL, cuneus. Lateral occipital gyrus, SPL, STG) and frontal (SFG), on both hemispheres; the left LOG with left SFG, MFG, STG, SMAR, and right IFG (all subdivision); the right PHG with temporal (left TP and STG), parietal (left precuneus, precentral) and occipitotemporal (right enthorinal, fusiform, inferior temporal) regions, mainly.

For PD-noVH patients, we found an overall greater significant of inter-regional correlation coefficients in the left and right pericallosal area, in the right lingual gyrus, and in the right superior frontal gyrus.

**Figure S8.1.**
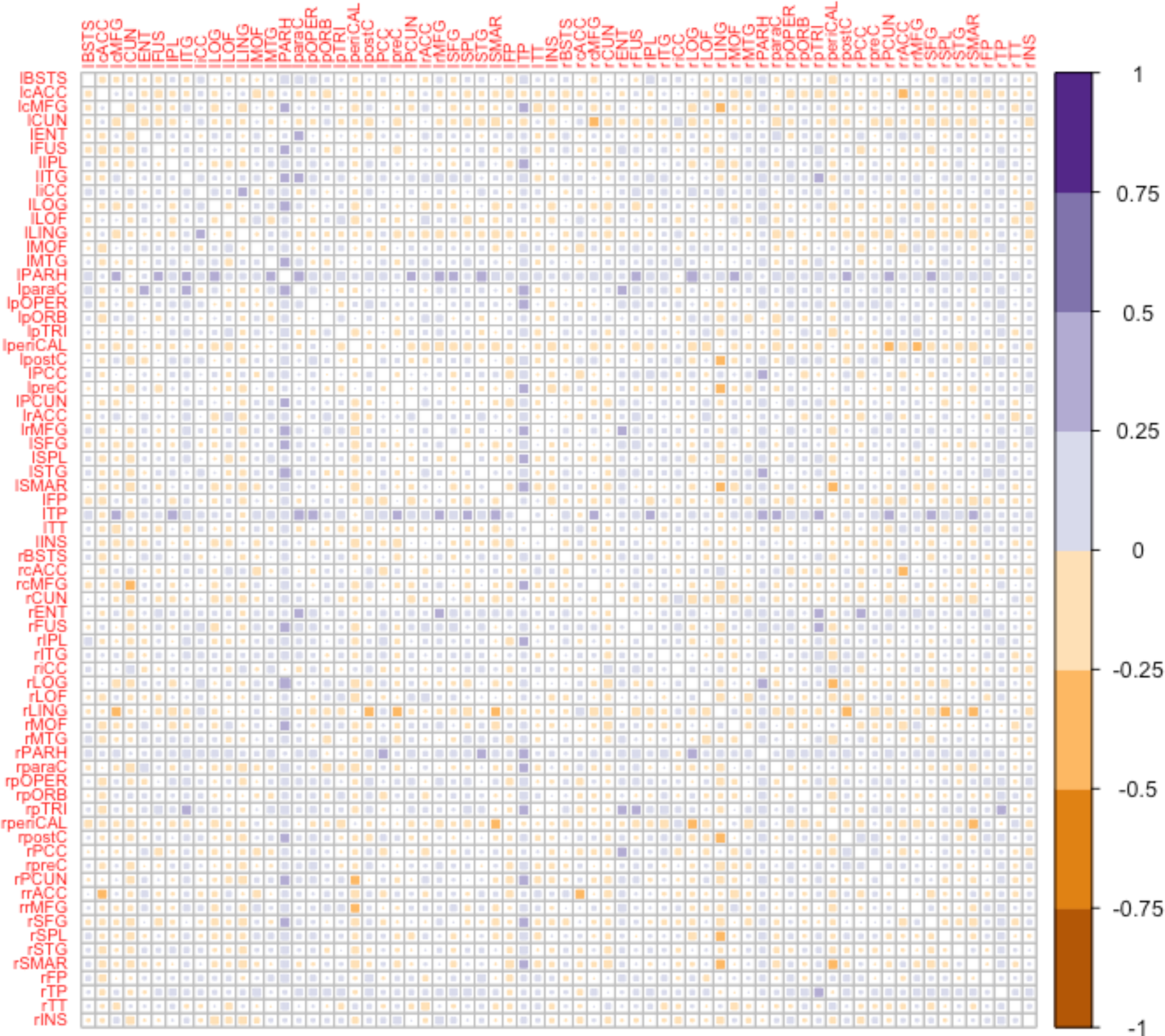
Correlogram showing the difference in inter-regional correlation coefficients in PD–VH vs. PD-noVH patients for cortical thickness. The yellow squares indicate inter-regional coefficients that were greater for PD-noVH; of those, only some were significant, mainly those in the left and right pericallosal column, in the right lingual column. For significant differences in inter-regional coefficients (z score) a.csv attachment will be made available upon publication on https://osf.io/fv2k7/registrations.)

The same procedure was carried out for *surface area*. However, we did not find any region where the correlation coefficient was greater for PD-noVH than for PD-VH. As for cortical thickness, regions with significant regions with the most inter-regional correlation with other regions were the left caudal MFG, left cuneus, left fusiform, right temporal transverse, right IFG triangualris, bilateral IFG opercularis and insula, IPL, SFG.

**Figure S8.2.**
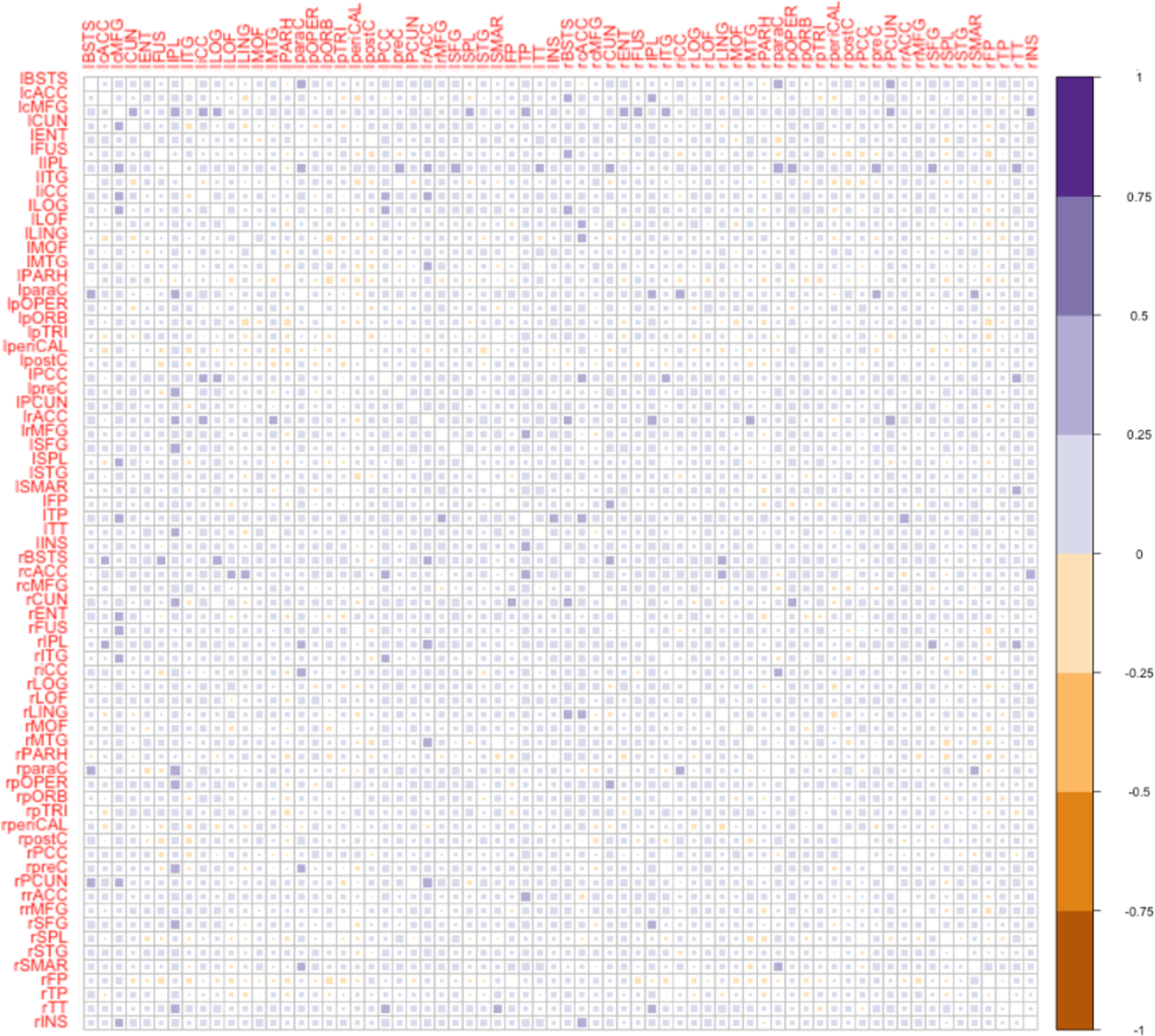
Correlogram showing the difference in inter-regional correlation coefficients in PD–VH vs PD-noVH patients for cortical surface area. The red squares indicate inter-regional coefficients that were greater for PD-noVH, with only the inter-regional coefficients for the left IFG orbitalis with the left PGH, left lingual gyrus and left frontopolar area were significant. (For significant differences in inter-regional coefficients (z score) a .csv attachment will be made available upon publication on https://osf.io/fv2k7/registrations.)

**Figure S8.3.**
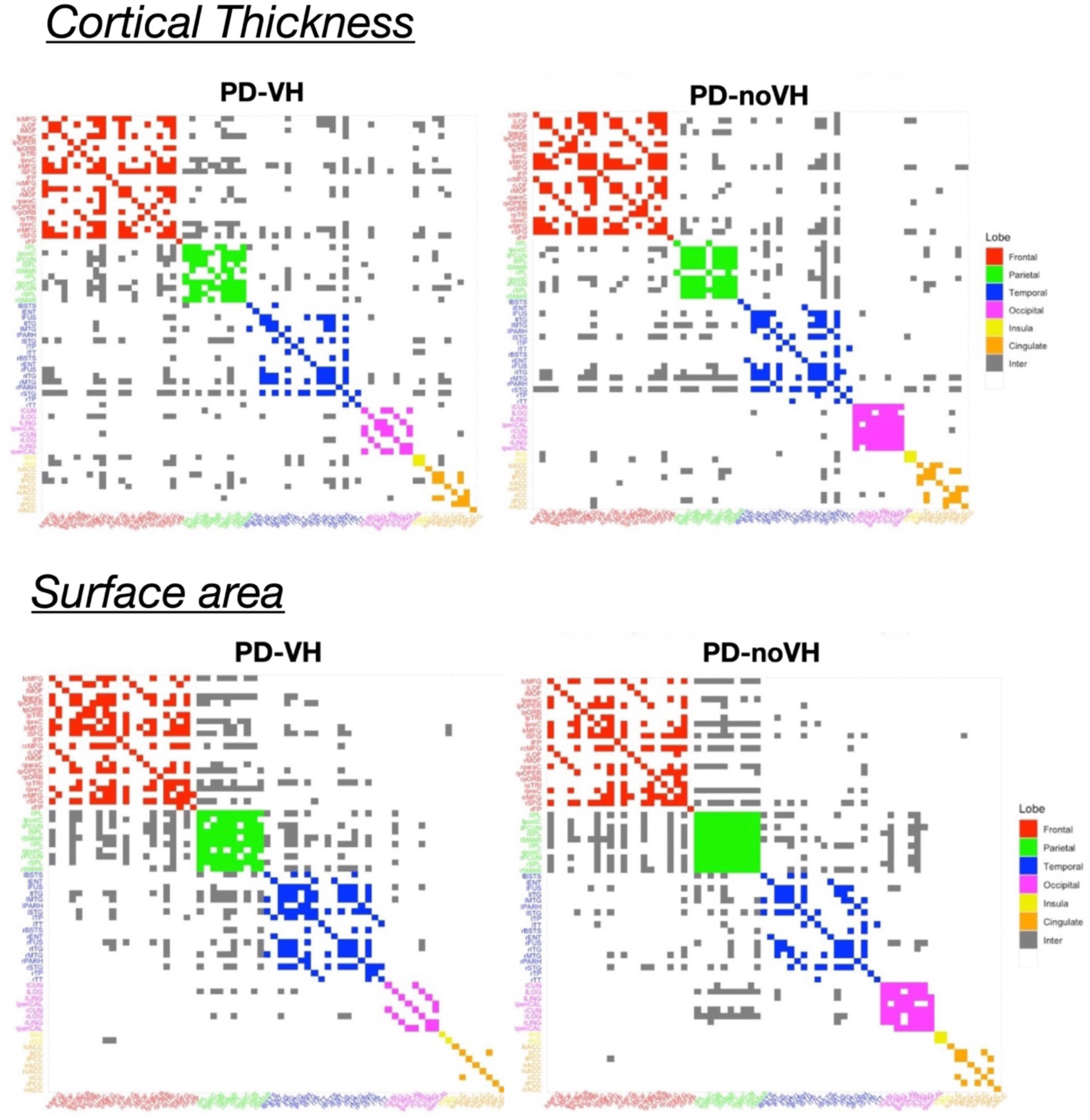
Community plot by lobe for cortical thickness (top row), surface area (bottom row). Legend: Red = frontal: bilateral caudal middle frontal gyrus, lateral orbital frontal gyrus, middle orbital frontal gyrus, paracentral gyrus, inferior frontal gyrus orbital, opercularis, triangular, precentral gyrus, middle frontal gyrus, superior frontal gyrus, frontopolar. Green = parietal: bilateral inferior parietal lobe, postcentral gyrus, precuneus, superior parietal lobe, supra marginal gyrus. Blue = temporal: banks superior temporal, enthorinal, fusiform gyrus, inferior temporal gyrus, middle temporal gyrus, parahippocampal gyrus, superior temporal gyrus,temporoparietal, temporal transverse. Pink = occipital: cuneus, lateral occipital gyrus, lingual gyrus, pericallosal. Yellow = Insula. Orange = cingulate: anter

